# Long-read sequencing of 945 Han individuals identifies novel structural variants associated with phenotypic diversity and disease susceptibility

**DOI:** 10.1101/2024.03.21.24304654

**Authors:** Jiao Gong, Huiru Sun, Kaiyuan Wang, Yanhui Zhao, Yechao Huang, Qinsheng Chen, Hui Qiao, Yang Gao, Jialin Zhao, Yunchao Ling, Ruifang Cao, Jingze Tan, Qi Wang, Yanyun Ma, Jing Li, Jingchun Luo, Sijia Wang, Jiucun Wang, Guoqing Zhang, Shuhua Xu, Feng Qian, Fang Zhou, Huiru Tang, Dali Li, Fritz J Sedlazeck, Li Jin, Yuting Guan, Shaohua Fan

**Author notes:** These authors contributed equally to this work (Jiao Gong, Huiru Sun, Kaiyuan Wang).

## Abstract

Genomic structural variants (SVs) are a major source of genetic diversity in humans. Although numerous studies explore SV diversity across global populations and their potential impacts^1–3^, validation using model systems are needed to confirm the reported genotype-phenotype associations. Here, through long-read sequencing of 945 Han Chinese genomes, we identify 111,288 SVs, including 24.56% unreported variants, many predicted to be functionally important. Our analysis unveils the multifaceted origins of these SVs within the Han population, with approximately 24% emerging at the common ancestor of modern humans. By integrating human population-level phenotypic, metabolic, and immunologic data and two humanized mouse models, we demonstrate the causal roles of two SVs: one SV that emerges at the common ancestor of modern human and Neanderthal and Denisova in *GSDMD* for bone density and one modern-human-specific SV in *WWP2* impacting height, weight, fat, craniofacial phenotypes, and innate immunity. Some of these phenotypes were previously unreported and irreproducible phenotypes in mouse knockout experiments. Our results suggest that the SV in *GSDMD* could serve as a rapid and cost-effective predictive biomarker for evaluating GSDMD-mediated pyroptosis in multiple organ injuries, including cisplatin-induced acute kidney injury. While initially identified in the Han, the functional conservation from human to mouse, signals of positive natural selection specifically in non-African populations including the Han, and associations with multiple disease risks, suggest that both SVs likely influence local adaptation, phenotypic diversity, and disease susceptibility across many non-African populations.

## Main

Structural variants (SVs) are genomic alterations ≥50 base pairs (bps) in length that result from duplications, deletions, insertions, inversions, and translocations^1–3^. The medical relevance of SV in the human genome has long been recognized, dating back to the advent of karyotyping techniques which enabled the identification of large chromosomal aneuploidies (∼3 Mb or more) and heteromorphisms leading to various genetic syndromes (reviewed in^4–6)^. The advances in cytogenetic (such as fluorescence in situ hybridization, FISH) and molecular techniques, such as bacterial artificial chromosome (BAC) array-comparative genomic hybridization (array-CGH), representational oligonucleotide microarray analysis (ROMA) and single nucleotide polymorphism (SNP) array, have enabled finer-scale genome-wide investigation of SV across populations^7,8^. These approaches allow the discovery of intermediate-size structural variants, ranging from kilobases to megabases in size, that are common among phenotypically normal individuals^7,8^. Their findings also suggest that SV may account for an equal or even greater amount of genetic diversity in the human genome compared to single nucleotide polymorphisms (SNPs) which were previously thought to be the primary source of human phenotypic diversity and disease susceptibility differences^7,8^. While FISH, array-CGH, and ROMA have made significant contributions to SV detection, they have inherent limitations in resolution, breakpoint, and complex SV detection^4,5,9–11^.

Short-read sequencing (SRS) technologies and development of the SV detection algorithms have greatly improved the identification and delineation of SVs and their precise breakpoints^2,12–17^. The 1000 Genomes Project was a pioneer SRS-based study that investigated the SV diversity of 2,504 individuals in global populations, albeit at low coverage (on average ∼7X)^2,17^. Subsequently, numerous studies have utilized SRS-based technologies to investigate SV diversity in global populations^12–16^. These studies have significantly deepened our understanding of SV diversity and their functional impacts in modern humans and suggested that the germline SVs affect an order of magnitude more nucleotides than SNPs^17^. However, they likely missed a large number of SVs, owing to limitations of SV calling based on SRS data (e.g., short read length, and difficulty in sequencing highly repetitive regions and regions with extreme GC content^18,19^).

The introduction of long-read single molecular sequencing (LRS) technologies, including Pacific Biosciences (PacBio) and Oxford Nanopore Technologies (ONT), has enabled increasingly comprehensive detection of SVs^18,19^. With an average read length >10 kb, and sometimes extending to several million bps, LRS can comprehensively cover SVs in entire regions of SVs^18,19^. For example, a survey of 32 individuals from 25 diverse populations (ethnicities) using LRS identified 107,136 SVs, among which only 29.6% could be detected using the SRS data for the same samples^3^. Notably, an increasing number of studies have made great strides in investigating SVs using LRS technologies, which has significantly enhanced our understanding of SV diversity^1,3,20–27^ and their potential associations with human phenotypic variations^3,22,23^, local adaptation^24–26^ and disease susceptibility^3,21–23^. However, functional validation using model systems is still needed to confirm the reported genotype-phenotype association signals attributed to SVs. A lack of understanding of the causal involvement of SVs in human local adaptation, phenotypic diversity, and disease susceptibility differences will not only hinder our understanding of human evolutionary history and phenotypic diversity but also constrain the development of personalized medicine.

In this study, we constructed a long-read-based SV catalog of 945 Han Chinese samples. We conducted extensive orthogonal validations, characterized the frequency distribution and location of SVs, and identified a significant number of previously unreported variants. Moreover, our analyses suggested a multifaceted origins for the SVs in our cohort and some of them can be traced back to chimpanzee. Based on the analyses of population-level multi-omics data from an independent Han Chinese cohort as well as two humanized mouse models, we identified two causal SVs for human phenotypic diversity and disease susceptibility difference. These models also allowed us to identify phenotypes that were previously reported in mouse gene knockout experiments that did not replicate in humans or humanized mice. We also studied the origin of the causal variants and revealed their differentiated selective pressures among ancestrally diverse populations based on comparison to the data from the chimpanzee reference genome, four archaic hominids, and 3,201 diverse modern humans from the 1000 Genomes Project.

### Generation of a long-read-based SV dataset of the Han population

We conducted whole-genome sequencing on 945 samples of Han ancestry using ONT with the PromethION platform. After base calling and quality control of the raw data (**Methods**), we obtained an average of 50 Gb of clean reads per individual, equating to a mean sequencing depth of 17X. We employed NGMLR^28^ to align the clean reads to the human reference genome GRCh38 with parameters specifically tailored for ONT reads to ensure precision and minimize potential biases. Our SV detection strategy involved a joint calling approach across the population. Initially, SVs were identified in each sample utilizing cuteSV^29^. Subsequently, SVs whose breakpoints within 500 bps of each other across different individuals were merged using SURVIVOR^30^ to avoid double counting of SVs at similar positions. Using this consolidated SV set, we re-genotyped each sample employing LRcaller^23^ and remerged the SVs, thereby obtaining a comprehensive, population-level SV dataset (**Methods**).

We detected 111,288 SVs, encompassing 42,300 insertions, 49,518 deletions, 13,503 duplications, 5,595 inversions, and 372 translocations (**Fig. 1a**). In agreement with prior studies^1,23^, we observed distinct peaks at sizes of approximately 300, 2,500, and 6,000 bps (**Fig. 1b**). These peaks likely correspond to the retrotransposition activities of Alu, SINE-VNTR-Alu (SVA), and LINE elements, respectively^2,23,31^. On average, we identified 23,729 SVs per sample, with counts ranging from 22,276 to 26,763, consistent with previous studies based on LRS technologies^1,3,23,32^ (**Fig. 1c**). Additionally, SVs collectively impacted an average of 17.83 Mb of genomic sequences per individual, with insertions and deletions accounting for 82.68% of the total sequences.

**Fig. 1:**
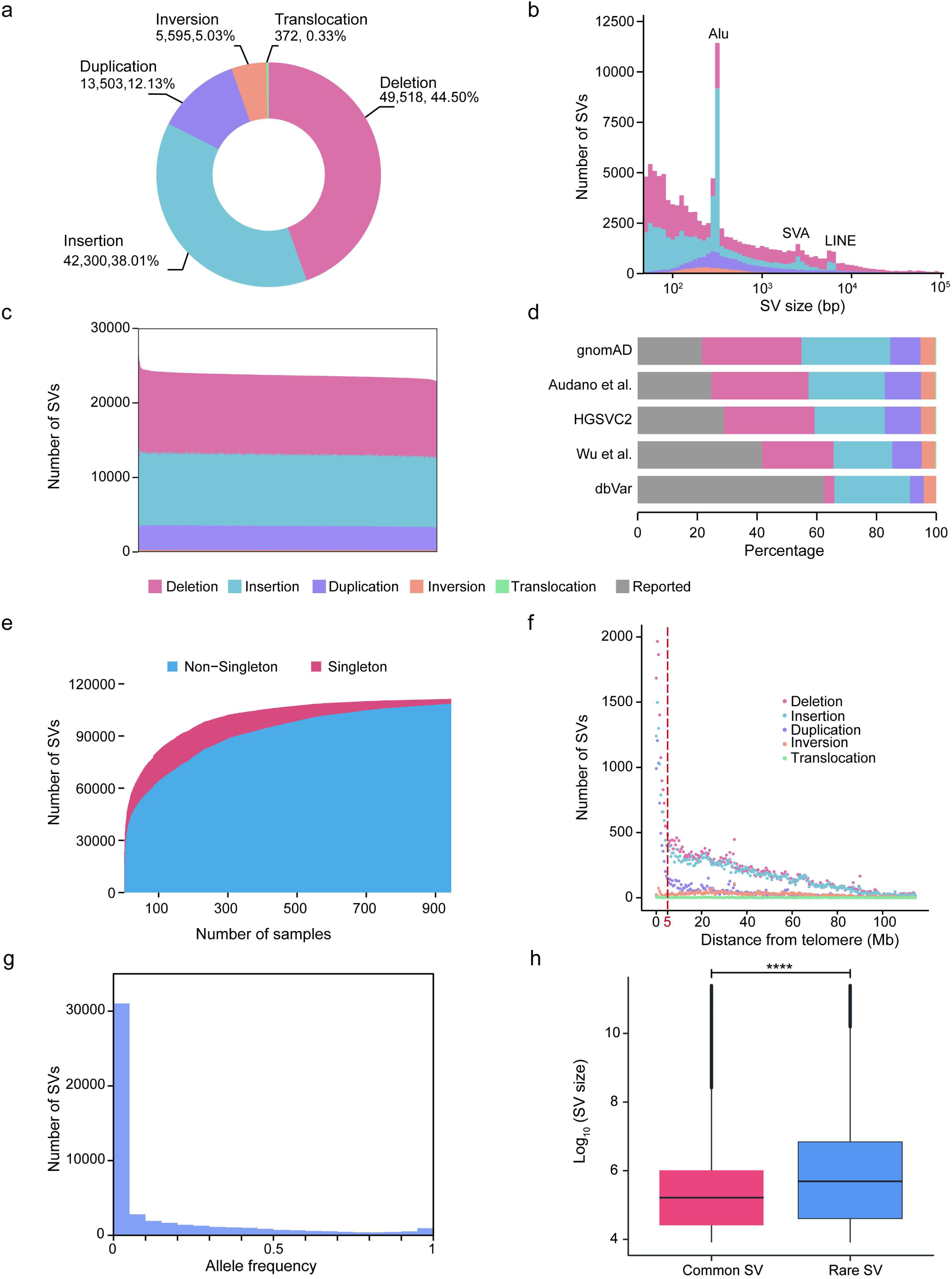
Summary of the SVs across the Han population. **a**, A total of 111,288 SVs, encompassing 42,300 insertions, 49,518 deletions, 13,503 duplications, 5,595 inversions, and 372 translocations, were identified in the present study. **b,** Distribution of SV size. Three peaks at sizes of approximately 300, 2,500, and 6,000 bps correspond to the retro-transposition activities of Alu, SINE-VNTR-Alu (SVA), and LINE elements, respectively. The X- and Y-axes indicate the size and number of SVs, respectively. **c,** Number of SVs per sample. Each bar represents a sample. We used color to indicate different types of SV per sample. **d,** Number of reported and unreported SV according to comparisons with different studies. **e,** Number of singleton and non-singleton SVs across samples. The X- and Y-axes indicate the number of samples and the number of singleton and non-singleton SVs when adding new samples, respectively. The decrease in singletons reached a plateau at a sample size >700, indicating that the present study captured most of the SV diversity in the Han population. **f,** A four-fold higher SV density was observed in subtelomeric versus other genomic regions (binned at 500 kb). **g,** A negative correlation was observed between allele frequency and the number of SVs. Rare SVs (minor allele frequency <0.05) comprise 64.22% of the SVs in the present study. The X- and Y-axes indicate the allele frequency and number of SVs, respectively. **h,** The sizes of rare SVs with minor allele frequencies <0.05 are significantly greater than those of common SVs. The X-axis indicates rare and common SVs, and the Y-axis shows the SV size. *P* value was calculated by a two-sided Wilcoxon rank sum test, \*\*\*\**p* <0.0001.

### Confirmation that the SV dataset is high-quality

We assessed the false discovery rate in our dataset using orthogonal methods (polymerase chain reaction (PCR) and Sanger sequencing) and comparisons to SVs with allele frequency (AF) >0.5 in global populations. We first employed PCR and Sanger sequencing to validate a set of 91 SVs, encompassing 39 insertions, 42 deletions, four inversions, and six duplications. We achieved a 100% validation rate for all 91 SVs, ranging from 53 to 81,592 bps (**Supplementary Fig. 1-2, Supplementary Table 1**). Additionally, we adopted a method previously developed in a study on SV diversity in Icelanders^23^, allowing us to estimate false-positive and false-negative rate boundaries of approximately 3.11–3.97% and 3.13–3.54%, respectively, based on comparisons with high-frequency SVs (AF >0.5) in global populations^1–3^ (**Methods**). Collectively, our analyses strongly indicate that a substantial proportion of the SVs within our dataset represent genuine genomic polymorphisms within the Han Chinese population.

### Discovery of a large number of unreported SVs

We uncovered a substantial number of previously unreported SVs within our dataset (**Fig. 1d**). Using a threshold of reciprocal overlap rate of ≥50%, we identified 87,308 (78.45%) SVs that were not reported in the gnomAD project^31^, a high-coverage (average depth of coverage of 32X) SRS dataset of 14,891 global samples (**Fig. 1d**). In addition, 83,845 (75.34%) and 79,153 (71.12%) of the SVs in our dataset were not reported in two LRS- based studies of SV diversity of global samples^1,3^ (**Fig. 1d**). Furthermore, 64,936 (58.35%) SVs in the present study were not found in a study of SV diversity based on 405 Chinese samples using LRS technology^22^ (**Fig. 1d**). Finally, 41,870 (37.62%) SVs have not been reported in the dbVar database^33^, which catalogs the SVs from a collection of 219 studies utilizing multiple platforms. Overall, 24.56% (27,333/111,288) of the SVs in the present study had not been documented in previous studies (**Fig. 1d**). Among the unreported SVs, we identified 780 SVs were located in the exonic regions of 670 genes, among which 182 SVs were predicted to cause open reading frame shifts, as well as 2,714 in transcription factor binding sites, 1,836 in strong enhancers, 614 in insulators based on the annotations of various cell lines from the ENCODE project^34^ using ANNOVAR^35^ (**Methods**). As expected, we found that the Afs of unreported SVs (median AF = 0.003) were significantly lower (***P* value <2.2e-16, two-sided Wilcoxon rank sum test**) than the AFs of the reported SVs (median AF = 0.02).

We computed the count of unique SVs (exclusively found in a single sample) across various sample sizes to assess the representativeness of identified SVs in the Han population^1^ (**Fig. 1e**). Notably, as the sample size expanded, the number of singleton SVs decreased continuously until reaching a plateau after ∼700 samples (**Fig. 1e**). This observation indicates that the sample size employed in our study effectively encompasses the majority of common SVs within the genomes of the Han population.

### Identification of the genomic features of SVs

We investigated the distribution of SVs across the genome and observed a four-fold higher SV density within the subtelomeric regions (within 5 Mb from the telomere regions) than in other regions (**Fig. 1f**), potentially related to the increased double-strand breakage^1,36^ and recombination^1,37,38^ rates and biased gene conversion^39^ in subtelomeric regions. In addition, 79.72% of the SV breakpoints overlapped with repeat elements, such as tandem repeats (17.84%) and interspersed repeats (57.51%), indicating the crucial role of repeat elements in SV formation^40^.

The average size of the SVs was 1,754 bps, ranging from 50 to 99,743 bps. In addition, 79.22% (87,868/110,916) of the SVs were <1,000 bps. When comparing the sizes of different types of SVs, we observed that insertions (average length = 521 bps) were shorter on average than deletions (average length = 2,340 bps), inversions (average length = 3,251 bps), and duplications (average length = 2,848 bps).

We noted a negative correlation between allele frequency and the number of SVs (**Fig. 1g**), aligning with the allele frequency distribution observed for single nucleotide variations in human genomes^41,42^. The rare (minor allele frequency [MAF] <5%) and common (MAF ≥5%) SVs comprised 64.22% and 35.78% of all identified SVs in this study, respectively. The sizes of rare SVs were significantly greater (***P* value <2.2e-16, two-sided Wilcoxon rank sum test**) than those of common SVs (**Fig. 1h**), which is in agreement with observations from previous studies^2,43,44^. Among the common SVs, 2,917 SVs, including 1,183 deletions, 237 duplications, 1,472 insertions, 22 inversions, and three translocations, were fixed (AF = 1) or nearly fixed (AF >0.9) in the population. After excluding three translocations that cannot be genotyped, we genotyped the rest of the high-frequency SVs using the high-coverage (30X) SRS data of 2,503 samples across 26 populations in the 1000 Genomes Project^45^ using Paragraph^46^. We observed that 96.51% of the SVs have allele frequency >5% in at least 13 populations in the 1000 Genomes project, suggesting that the reference genome carries minor alleles at these loci across the global populations (**Methods**).

### The origin of the SVs in the Han

Our analyses suggest multifaceted origins for the SVs in our cohort based on the genotyping of 110,863 SVs (after excluding 372 translocations and 53 SVs containing non-A/G/T/C bases in their alternative sequences in the human reference genome) in 2,503 unrelated high-coverage SRS samples in the 1000 Genomes Project using Paragraph^46^ and a comparison with the SVs obtained based on LRS data of a chimpanzee genome^47^ (**Methods**).

Our analyses of SVs in the Han Chinese population reveal insights into the evolutionary origins and dynamics of genomic variation (**Fig. 2a**). We find that approximately 2% (2,273 SVs affecting 807 genes) of SVs are ancient polymorphisms shared between humans and chimpanzees, indicating that they originated before the divergence of these species (**Fig. 2b**). An additional 4% (4,140 affecting 1,395 genes) likely emerged in the common ancestor of modern humans, Neanderthals, and Denisovans (**Fig. 2b**). Notably, about 24% (26,417 affecting 5,990 genes) of the identified SVs appear to be modern human-specific, as they are present across diverse populations around the world but absent from archaic hominin genomes. These variants may have played important functional roles in the more recent evolution of anatomical and physiological traits unique to Homo sapiens (**Fig. 2b**).

**Fig. 2:**
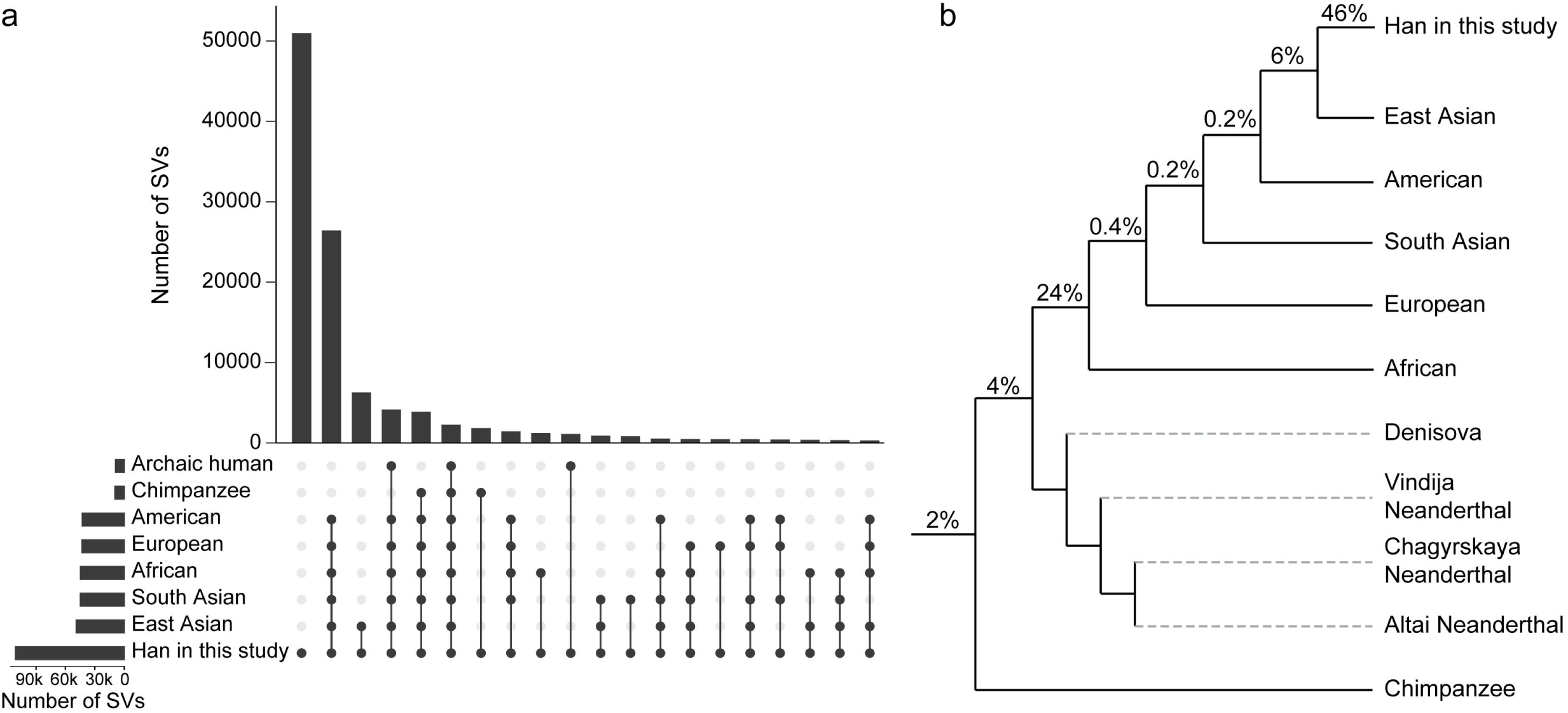
The multifaceted origins of SVs in the Han. **a**, UpSet plot of the numbers of SVs shared among chimpanzees, archaic, and modern humans. We displayed the top 20 with the highest number of SVs across groups. **b,** The evolution of SV in a phylogenetic context. The percentages on each branch indicate the proportion of SVs in the Han lineage estimated to have arisen during the given evolutionary period. The phylogeny is based on the results of previous studies^2,119–124^.

Within modern human groups, we also uncover SVs that trace their origins to more recent common ancestors at continental and sub-continental scales (**Fig. 2b**). For example, 0.4% (458 affecting 221 genes) evolved at the common ancestors of Europeans, Americans, and Asians, 0.2% (181 affecting 99 genes) evolved at the common ancestors of Americans and Asians, and ∼6% (6,287 affecting 3,195 genes) were evolved at the common ancestors of East Asians including the Han in the present study (**Fig. 2b**). Intriguingly, a small proportion (∼0.1%, 59 affecting 23 genes) of SVs show signatures of introgression from Neanderthals or Denisovans, indicating occasional gene flow from now-extinct hominin populations into ancestors of present-day Asians (**Fig. 2a**). Finally, approximately 46% (50,960) of identified SVs appear specific to our Han cohort and likely represent de novo mutations (**Fig. 2b**). The majority of these variants are rare, with a median allele frequency of ∼0.004, within the cohort.

### Characterization of a deletion shared by modern and archaic humans in *GSDMD* associated with acute kidney injury, bone density, and levels of sphingomyelin and phosphatidylcholine

To explore the potential functional impacts of the SVs, we performed a gene-based annotation based on NCBI RefSeq annotations (as of August 17, 2020) using AnnotSV^48–50^. A total of 51.24% (57,026/111,288) and 44.13% (49,110/111,288) of the SVs were located in intergenic and intronic regions, respectively, while 4.63% (5,152/111,288) of the SVs overlapped with at least one exon of 3,326 genes. Among the exonic SVs, 28.92% (1,490/5,152) can potentially disrupt gene function through open reading frame shifts. Based on a gene-based gene ontology enrichment analysis using the Database for Annotation, Visualization and Integrated Discovery (DAVID, updated on October 11, 2023)^51,52^ (**Methods**), we found that the genes with SVs located in their exonic regions were significantly enriched in the pathways involved in regulation of keratinization (**FDR-adjusted *P* value = 0.008**), transcription (**FDR-adjusted *P* value = 0.009**), and metal ion binding (**FDR-adjusted *P* value = 0.06**) (**Supplementary Table 2**).

Regarding common SVs (MAF ≥5%), the SV location pattern was similar to that of all SVs, with 3.75% (1,492/39,817) of SVs located in the exonic regions of 814 genes. Gene set enrichment analysis conducted using DAVID indicated significant involvement of the genes with common SVs located in their exonic regions in pathways related to immunity (immune response, **FDR-adjusted *P* value = 0.01**; defense response to Gram-negative bacterium, **FDR-adjusted *P* value = 0.04**), keratinization (**FDR-adjusted *P* value = 0.01**), gas transport (carbon dioxide transport, **FDR-adjusted *P* value = 0.08**; oxygen transport, **FDR-adjusted *P* value = 0.097**), sensory perception of taste (**FDR-adjusted *P* value = 0.08**), and digestion (**FDR-adjusted *P* value = 0.097**) (**Supplementary Table 3**). Among the immune-related genes, we identified a 2,175-bps deletion (chr8:143,551,891‒143,554,066, GRCh38) with an allele frequency equal to 43% that eliminated the first exon of the longest isoform of *GSDMD* (NM_001166237.1) (**Fig. 3a, b**). The deleted region is enriched with signals of histone modifications and transcription factor binding sites (**Fig. 3b**). It was also predicted to function as an enhancer to different isoforms of *GSDMD* (**Fig. 3b**) according to the dual elite annotation from the GeneHancer database^53^, which consolidates high-confidence enhancer-gene associations from multiple sources, including Hi-C, expression quantitative trait locus, and enhancer information. *GSDMD* encodes a pore-forming effector protein that facilitates inflammatory cell death, also known as pyroptosis^54^.

**Fig. 3:**
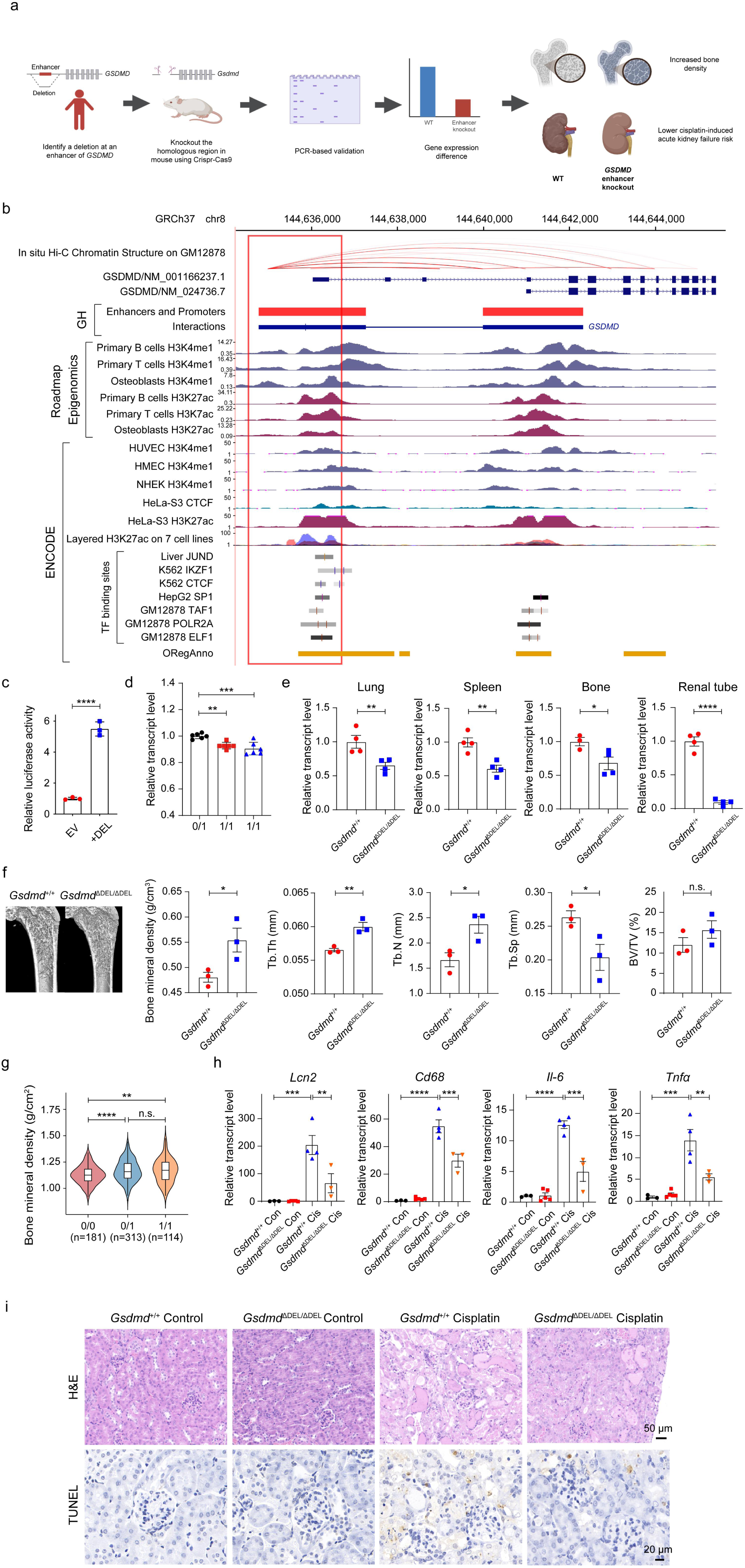
A deletion at the *GSDMD* locus is associated with bone density and acute kidney injury risk in mice and humans. **a**, Schematic of the study of the functional impacts of the deletion at the *GSDMD* locus. A targeted deletion was introduced in the mouse genome using CRISPR/Cas9 technology and validated the deletion using PCR. We examined the impacts of the target deletion on bone density and acute kidney injury risk. **b,** Genomic context around the deletion (indicated by the red box) at the *GSDMD* locus, including the Hi-C interaction sourced from Rao et al., 2014^113^, annotations for promoters and enhancers from the GeneHancer database, H3K4me1 and H3K27ac ChIP-seq tracks for primary B cells, primary T cells and osteoblasts from the Roadmap Epigenomics dataset, H3K4me1, H3K27ac or CTCF ChIP-seq tracks for HUVEC, HMEC, NHEK, HeLa-S3, a layered H3K27ac track on 7 cell lines (GM12878, H1-hESC, HSMM, HUVEC, K562, NHEK and NHLF), and transcription factor binding sites (TFBS) tracks for Liver, K562, HepG2 and GM12878 from ENCODE data set. All the annotations were visualized in the UCSC Genome Browser. **c,** Luciferase reporter assays of the sequence affected by the deletion. The depleted sequence is significantly associated with elevated enhancer activity compared with that of the empty vector. The dots represent biological replicates, of which three were conducted. **d,** Human carriers of the homozygous deletion have significantly lower *GSDMD* expression than heterozygous carriers, as demonstrated by RT-qPCR. Each bar represents a sample, and each dot is a technical replicate. We conducted six technical replicates per sample. **e,** Knocking out the homologous region in mice causes significant decreases in *Gsdmd* expression in lung, spleen, bone, and renal tubule cells. **f,** Knocking out the homologous region in mice causes a significant increase in traits related to bone density. We measured the bone volume/tissue volume (BV/TV) of proximal tibial trabeculae, trabecular number (Tb. N), trabecular thickness (Tb. Th), and trabecular separation (Tb. Sp) as determined by microCT in 7-week-old *Gsdmd* enhancer KO mice and their littermates. **g,** Human carriers of the deletion exhibit a significant increase in bone density compared to noncarriers. **h,** *Gsdmd* enhancer KO mice exhibit less severe acute kidney injury than wild-type littermates after cisplatin administration according to the expression of four acute kidney injury markers, *Lcn2*, *Cd68*, *Il-6,* and *Tnfα*. (**i**) Representative images of H&E- and TUNEL-stained kidney sections from WT littermates and *Gsdmd* homozygous enhancer KO mice following saline or cisplatin injection. Scale bar: 50 μm (HE); 20 μm (TUNEL). All data are represented as the mean ± SEM. *P* values were calculated by one-way ANOVA for d, g and h, and by two-tailed *t-*test for c, e and f. \**p* <0.05, \*\**p* <0.01, \*\*\**p* <0.001, \*\*\*\**p* <0.0001, n.s. non-significant.

Luciferase reporter assays were conducted in HeLa cells to validate the enhancer activity of the region affected by the deletion (**Methods**). Our experiments demonstrated a significant increase in luciferase signal (***P* value <0.0001, two-tailed *t-*test**) when using plasmids containing the sequence of the deleted region compared to empty vectors after normalization. This finding indicates that the region affected by the deletion is likely to function as an enhancer (**Fig. 3c**). In agreement, the homozygous carriers of the deletion exhibited significantly reduced *GSDMD* expression (***P* value <0.01, one-way analysis of variance (ANOVA)**) compared to heterozygous carriers, as determined by RT-qPCR (**Fig. 3d**). In summary, our findings suggest that the deletion depletes the first exon of the longest *GSDMD* isoform, resulting in the downregulation of *GSDMD* through the disruption of their enhancer activity.

To investigate the functional impacts of the deletion at the *GSDMD* locus *in vivo*, we employed CRISPR/Cas9 technology to generate a targeted deletion of a 1,410-bps homologous region in the mouse genome (**Fig. 3a; Methods**). The knockout (KO) mice were born at the expected Mendelian ratio and grew normally. In line with findings observed in humans (**Fig. 3d**), RT-qPCR analyses revealed that the excision of the homologous region in mice resulted in significant reductions (***P* value <0.05, two-tailed *t-*test**) in *Gsdmd* expression levels compared to those in their wild-type (WT) littermates across a spectrum of tissues, encompassing the spleen, lung, bone, and renal tube (**Fig. 3e**). These results indicate that the deleted region is likely to act as an enhancer regulating *Gsdmd* expression in the mouse genome.

Notably, contrary to a decrease in bone density when knocking out *Gsdmd* gene in mice^55^, we observed the opposite effect in humanized mouse models and human carriers of the deletion. Humanized *Gsdmd* enhancer KO mice exhibited increases in BMD, trabecular thickness (Tb.Th), and trabecular number (Tb.N), along with reduced trabecular separation (Tb.Sp), relative to wild-type littermates (***P* value <0.05, two-tailed t-test, Fig. 3f**). However, no significant difference occurred in the bone volume/tissue volume (BV/TV) of proximal tibial trabeculae between the *Gsdmd* enhancer KO mice and wild-type littermates (**Fig. 3f**). Additionally, humans with the *Gsdmd* structural variant showed significantly increased bone mineral density (BMD) compared to those without the variant (***P* value <0.0001, one-way ANOVA, Fig. 3g**).

Our results suggest the SV in *GSDMD* could potentially serve as a biomarker to evaluate risk for multiple diseases. For example, we observed that introducing the human mutation into mouse model confers a protective effect against cisplatin-induced acute kidney injury, evidencing for significant decreases in the expression levels of four key acute kidney injury markers, namely, *Lcn2*, *Cd68*, *Il-6*, and *Tnfα*, in *Gsdmd* enhancer KO mice versus wild-type littermates after cisplatin administration (**Fig. 3h**). This observation was further substantiated by histological analysis, which demonstrated reduced tubular dilation and immune cell infiltration in the enhancer KO mice (**Fig. 3i**) and decreased cell death compared to their wild-type littermates (**Fig. 3i**).

We also observed the genotype-phenotype associations that have not been reported in previous *Gsdmd* KO mouse models. The carriers of the SV have significant increases of 13 phosphatidylcholines (***P* value < 0.05**, **Kruskal-Wallis test**) and one sphingomyelin (***P* value < 0.05, Kruskal-Wallis test**) than the non-carriers (**Supplementary Fig. 3**). Previous studies showed that sphingomyelin and phosphatidylcholine are not only crucial metabolites for maintaining cell normal function^56–59^ but also risk markers of multiple diseases. For example, the dysregulated sphingomyelin is an independent cardiovascular disease risk factor of atherogenesis^57^. Altered phosphatidylcholines levels have also been linked to obesity, atherosclerosis, and insulin resistance via effects on tissue composition^60^.

We next studied the evolution of the deletion at the *GSDMD* locus in humans based on the genotyping results of the high-coverage SRS data of 3,201 samples from 26 populations in the 1000 Genomes Project^45^ (**Fig. 4a and Supplementary Table 4**) and samples from four archaic humans^61–64^ (**Fig. 4b**) as well as the alignment of the human and chimpanzee reference genomes at this locus (**Methods**) and manual examination. Our analyses revealed that the deletion is absent in the chimpanzee reference genome (**Supplementary Fig. 4a**) and is present in genomes of three Neanderthals who are likely to be homozygous carriers and one Denisovan sample who is likely to be heterozygous carrier based on depth of coverage (**Fig. 4b**). In addition, it exhibits rarity (average AF = 0.03) within the African ancestry group but is common (average AF >0.1) across all non-African populations of the 1000 Genomes Project (**Fig. 4a**) except for the Peruvians with AF=0.02. Based on genotyping 102,118 SVs from a previous study of SV diversity in the global populations using LRS data^3^, we identified signatures of positive selection at this deletion in multiple non-African populations when calculating *F*ST (the fixation index) between non- African and African populations in the 1000 Genomes project (**Supplementary Fig. 5**). The deletion ranks in the top 2% of SVs exhibiting the highest *F*ST values between the Han and Luhya (**Supplementary Fig. 5; Methods**), aligning with the substantial differential frequencies observed between these populations (**Fig. 4a**). Collectively, our results suggest that this deletion is likely to have evolved in the common ancestor of modern humans (**Fig. 4b**), Neanderthals, and Denisovans and then underwent positive selection in non-African populations.

**Fig. 4:**
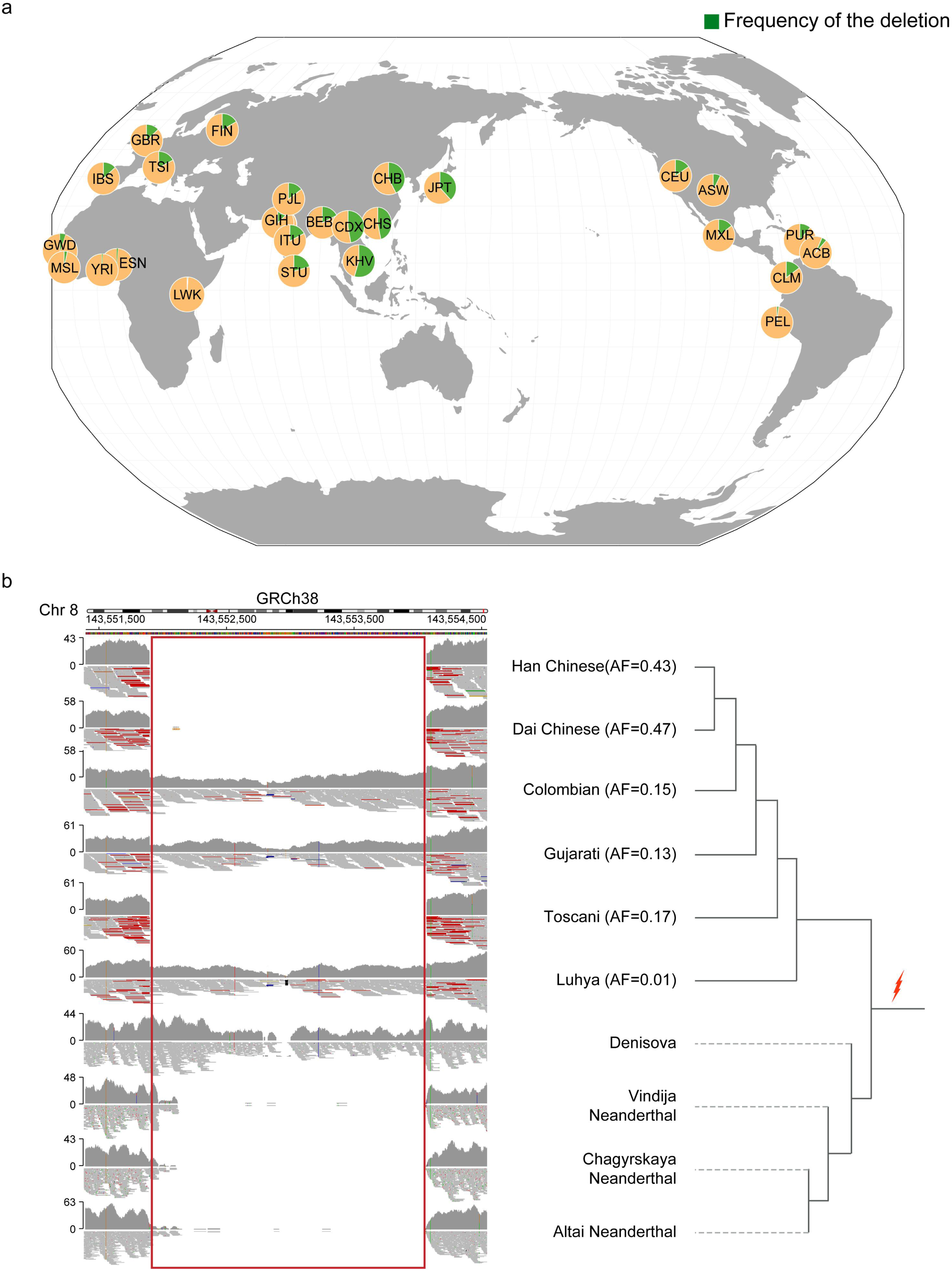
Evolution and demographic distribution of the deletion at the *GSDMD* locus. **a**, The allele frequency of the deletion (indicated in green) in the genomes of 3,201 samples from 26 populations in the 1000 Genomes Project^45^. In the pie charts, the frequencies of the reference and deletion alleles are depicted in yellow and green, respectively. **b**, The read mapping results around the deletion at the *GSDMD* locus in the genomes of four archaic humans and six representative modern human samples. The Han Chinese sample who carries a homozygous deletion is from the present study. We obtained the high-coverage SRS data of one Chinese-Dai individual in Xishuangbanna (CDX, ID: HG01046) who carries a homozygous deletion, one Gujarati Indians in Houston, TX (GIH, ID: NA20854) who carries a heterozygous deletion, one Toscani in Italy (TSI, ID: NA20581) sample who carries a homozygous deletion, one Colombian in Medellin, Colombia (CLM, ID: HG01261) who carries a heterozygous deletion, and one Luhya in Webuye, Kenya (LWK, ID: NA19323) who carries a heterozygous deletion. The read- mapping results suggest that the deletion (indicated by the red box) is present in the genomes of three Neanderthal samples who are likely to carry a homozygous deletion and one Denisovan sample who is likely to carry a heterozygous deletion. The phylogeny is based on the results of previous studies^2,119–123^. Due to its presence in the genomes of all modern human populations, three Neanderthals and one Denisovan, the deletion is likely to have emerged in the common ancestor of modern humans, Neanderthals, and Denisovans (highlighted by the red arrow).

### A novel complex SV specific to modern humans in *WWP2* is associated with shorter stature, increased body fat percentage, and enhanced immune response

We next investigated the regulatory potential of SVs, considering that 95.37% were located in intronic and intergenic regions, known to be enriched with regulatory elements. Our analysis revealed that 3.53% of the SVs entirely overlapped with 5,666 enhancers, as annotated in the GeneHancer database^53^. Regarding the common SVs (MAF ≥5%), 116 completely covered 158 enhancers regulating the expression of 277 protein-coding genes according to GeneHancer database annotations. A gene-based enrichment test using DAVID suggested that these genes are significantly involved in pathways related to the composition of the cytoplasm (cytoplasm, **FDR-adjusted *P* value = 0.08**, **Supplementary Table 5**).

We further explored the functional implications of the genes potentially regulated by the common SVs, harnessing the comprehensive resources of the Mouse Genome Informatics (MGI) website^65^. MGI catalogs comprehensive functional, phenotypic, and disease annotations of mouse genes, primarily derived from gene knockout experiments. Our investigation identified 121 genes within the mouse genome that are orthologous to the 277 genes regulated by SVs observed in humans according to human-mouse orthologous gene annotations in the MGD database^65^ (**Methods**). After excluding a union of 24 genes linked to lethality (23 genes) and had no discernible phenotypic consequences (two genes), as determined from knockout experiments in mice (**Methods**), the remaining 97 genes were demonstrated to likely exert influence across 833 distinct ontologies in mice. Notably, several top-ranking ontologies pertained to phenotypic attributes (such as decreased body weight, size, litter size, and postnatal growth retardation), physiological traits (such as impaired glucose tolerance and decreased susceptibility to diet-induced obesity), fertility (such as oligozoospermia, asthenozoospermia, and male infertility), and immune system physiology (**Supplementary Table 6**). For example, we identified a complex common SV (AF

= 0.28)—a 229-bps insertion (caused by the fusion of two SINE elements ∼4,225 bps apart from each other, **Fig. 5**) followed by a 354-bps deletion—located within the fourth intron of *WWP2* (NM_001270454.2) that is associated with multiple phenotypic (e.g., body weight and size and craniofacial traits) and immunological traits based on KO experiments in mice. However, the phenotypes reported in different studies are inconsistent. For example, one study observed reduced body length and weight along with abnormal teeth and craniofacial traits^66^. However, these phenotypes were not replicable in other studies^67,68^. These discrepancies impede the translation of phenotypes from mouse knockout experiments to humans and emphasize the need to validate model organism phenotypes using human genetic data. Additionally, while the dysregulation of *WWP2* has been implicated in the increased risks of cardiovascular diseases^69,70^, osteoarthritis^71^, and a novel oncogene (reviewed in^72^), no causal variant has yet been identified that regulates *WWP2* and its associated phenotypes.

**Fig. 5:**
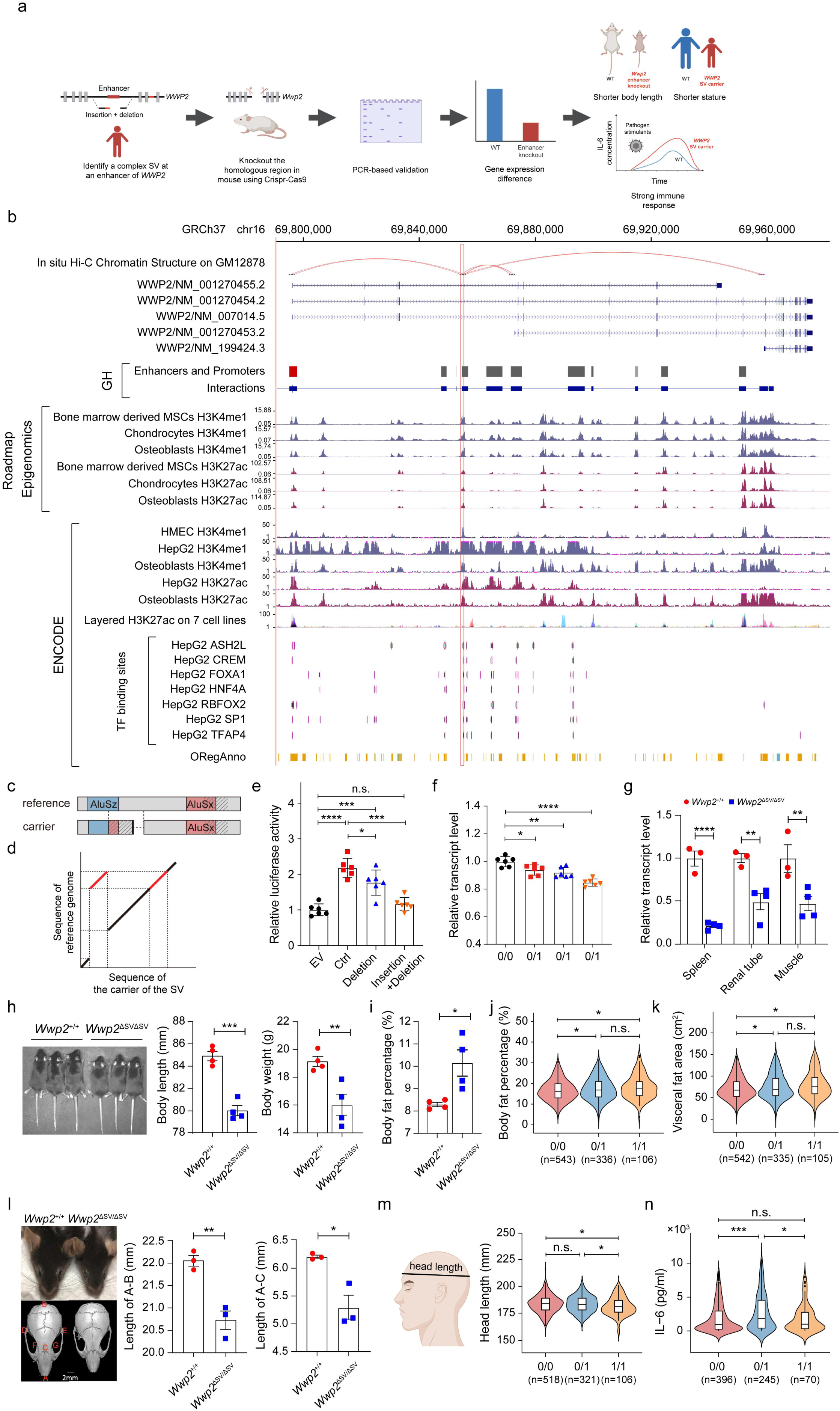
A complex SV at the *WWP2* locus is associated with height, body fat percentage, craniofacial traits, and immune response. **a**, Schematic of the study of the functional impacts of the SV at the *WWP2* locus. A targeted deletion was introduced in the mouse genome using CRISPR/Cas9 technology and validated the deletion using PCR. We examined the impacts of the target deletion on height and craniofacial traits as well as immune response. **b,** Genomic context around the 1,127-bps enhancer (indicated by the red box) at the *WWP2* locus, including Hi-C interaction sourced from Rao et al., 2014^113^, annotations for promoters and enhancers from the GeneHancer database, H3K4me1 and H3K27ac ChIP-seq tracks for Bone derived MSCs (E026), Mesenchymal stem cell derived chondrocyte (E049) and osteoblasts (E129) from the Roadmap Epigenomics dataset, H3K4me1, H3K27ac ChIP-seq tracks for HMEC, HepG2, Osteoblasts, a layered H3K27ac track on 7 cell lines (GM12878, H1- hESC, HSMM, HUVEC, K562, NHEK and NHLF), and transcription factor binding sites (TFBS) tracks for HepG2 from ENCODE data set. All the annotations were visualized in the UCSC Genome Browser. **c,** The SV consists of an insertion mediated by the fusion of two SINE elements followed by a 354-bps deletion. **d,** Dot plot of the sequences of the carriers of the SV (X-axis) and the human reference genome (Y-axis). **e,** SV significantly reduced the enhancer activity of the region according to the luciferase reporter assays. The dots represent biological replicates, of which six were conducted. **f,** Human carriers of the SV exhibit lower *WWP2* expression based on RT- qPCR than the non carriers. **g,** Knocking out the homologous sequence of the enhancer in mice led to significant reductions in *Wwp2* in multiple tissues, suggesting that the homologous sequence functioned as an enhancer of *Wwp2* in mice. **h-i,** Knocking out the homologous sequence of the enhancer led to a significant decrease in body length and weight (**h**), while it led to a significant increase in body fat percentage (BF%) (**i**) in mice. **j-k,** Human carriers of the SV exhibit higher BF% as well as larger visceral fat area than non-carriers. **l,** A comparison of craniofacial traits between *Wwp2* enhancer KO mice and wild-type littermates. **m,** Human carriers of SV exhibit significantly shorter head lengths than noncarriers, a trait homologous to skull length (distance between A and B in Fig. 5l). The X- and Y-axes indicate the different genotypes and head lengths, respectively. The numbers in the brackets are the number of samples. **n,** Human carriers of the heterozygous SV exhibit significantly higher levels of IL-6 than noncarriers when stimulated using flagellin, which is the principal component of bacterial flagella and is widely used as a potent activator of a broad range of cell types involved in innate and adaptive immunity. Due to a low sample size of the homozygous SV carriers, the difference IL-6 is not significant after flagellin stimulation. The mice were measured at five weeks of age. All data are represented as the mean ± SEM. *P* values were calculated by one-way ANOVA for e-f, two-way ANOVA for g, Kruskal-Wallis test for j, k m and n, and by two-tailed *t-*test for h, i and l. \**p* <0.05, \*\**p* <0.01, \*\*\**p* <0.001, \*\*\*\**p* <0.0001, n.s. non-significant.

We first validated this SV using PCR-based Sanger sequencing and local assembly of the long reads at this region (**Fig. 5a-d, Supplementary Fig. 6; Methods**). Our analyses further suggest that the SINE fusion event led to a false-positive detection of a ∼4.5 kb deletion signal in studies conducted with SRS and LRS technologies when the reads were mapped to this region (**Supplementary Fig. 6; Methods**).

The SV in *WWP2* overlapped a 1,127-bps annotated enhancer region (chr16:69,820,390‒69,821,516, GRCh38) that is likely to be active in multiple bone-related cells/tissues as well as breast, brain, duodenum and stomach tissues according to the annotations of the Roadmap Epigenomics Project^73^ and regulates the expressions of different isoforms of *WWP2* based on Hi-C data (**Fig. 5b and Supplementary Fig. 7**). The enhancer potential of the region was first validated using luciferase reporter assays. The plasmid containing the 1,127-bps sequence demonstrated significantly greater luciferase signal levels (***P* value <0.0001, one-way ANOVA**) than the empty vector after normalization (**Fig. 5e**). In addition, the presence of the SV, especially when both the insertion and the deletion were concurrently presented, significantly reduced luciferase signals (***P* value <0.0001, one-way ANOVA**), indicating that the complex SV disrupted the enhancer activity (**Fig. 5e**). Furthermore, RT-qPCR revealed that carriers of the SV exhibited a significant decrease in *WWP2* expression in the whole blood (***P* value <0.05, one-way ANOVA**) versus noncarriers (**Fig. 5f**).

The SV was associated with standing height in our cohort, individuals carrying this SV displayed a lower standing height (median height = 164 cm) than that of the noncarriers (median height = 165 cm), consistent with the reduced body length caused by knocking out of the *Wwp2* gene in mice^74,75^. While the difference was not statistically significant due to the relatively small sample size (n = 1,016) in the present study (***P* value = 0.28, one- way ANOVA**), we found that the SV is in complete linkage disequilibrium with rs8052428-T, which is ∼55 kb downstream to the SV, based on manual examination of the Pacbio HiFi data from a Chinese quartet family (http://chinese-quartet.org/)^76^. rs8052428-T is significantly associated with shorter sitting height (**Beta = -0.101, *P value* = 1.1E-29**) and standing height (**Beta = -0.132, *P value* = 1.0E-17**) in a prior genome-wide association study (GWAS) of height variation of 361,194 UK Biobank samples (http://www.nealelab.is/uk-biobank/). However, no causal variant has yet been identified at this locus. We observed that rs8052428 is located in the intronic region of *WWP2* and there is no enhancer annotation around the region (**Supplementary Fig. 8**). This supports the SV, rather than rs8052428, is the causal variant of height variation not only in the Han but also in samples of European ancestry.

To further understand the functional impacts of the SV, we depleted a 1,267-bps region, that is homologous to the SV-affected region based on the liftOver tool in the UCSC genome browser website^77^, in the mouse genome using CRISPR/Cas9 technology (**Methods**). The KO mice were born at the expected Mendelian ratio and grew normally. We observed significantly reduced *Wwp2* expression levels in the spleen, renal tube, and muscle tissues of the KO mice compared to those in the wild-type mice (**Fig. 5g, *P* value <0.01, two-way ANOVA**), confirming the homologous region is functional as an enhancer in the mouse genome. Furthermore, the *Wwp2* enhancer KO mice in the present study displayed significantly shorter body length and lower body weight (***P* value <0.01, two-tailed *t-* test**) than their wild-type littermates (**Fig. 5h**). Remarkably, the mice with the enhancer knocked out exhibited substantial rises in body fat percentages (BF%) (**Fig. 5i**, ***P* value <0.05, two-tailed t-test**) than the wild-type mice, which has not been observed in previous *Wwp2* KO mouse models. This result is further exemplified by observations in humans showing that carriers of the SV have significantly greater body fat percentage (**Fig. 5j**, ***P* value <0.05, Kruskal-Wallis test**) and larger visceral fat area (**Fig. 5k**, ***P* value <0.05, Kruskal-Wallis test**) than the non-carriers.

Deep phenotyping data in humans and introducing human mutations into mouse genomes have enabled the identification of the irreproducible and unreported phenotypes in previous mouse KO experiments. For instance, the abnormal craniofacial and dental changes reported in *Wwp2* knockout mice^66^ could not be replicated in our humanized mouse model and human carriers. Instead, we observed that the *Wwp2* enhancer KO mice exhibited a shorter skull length (**Fig. 5l. distance between A to B, *P* value <0.01, two-tailed *t-*test**), a more domed skull, and a shortened snout (**Fig. 5l. distance between A to C, *P* value <0.05, two-tailed *t-*test**) in comparison with the wild-type littermates using microCT. This was further exemplified by our human samples, as we only observed the head length (the distance between the glabella and opisthocranion), a homologous trait to skull length in mice, was significantly shorter in the SV carriers (***P* value <0.05, Kruskal-Wallis test**) than that in the noncarriers (**Fig. 5m**) but no other abnormal craniofacial or dental phenotypes were observed in human carriers.

Our results suggest that the SV in *WWP2* may yield an increased innate immune response to its carriers. While there was no significant difference in the IL-6 levels in the whole blood among the samples in the independent cohort (**Supplementary Fig. 9a**), we observed significant increases in IL-6 in response to flagellin stimulation, a subunit protein that polymerizes to form the filaments of bacterial flagella and a Toll-like receptor 5 agonist, in the carriers of the SV compared to that in the noncarriers (***P* value <0.05, Kruskal−Wallis test**) (**Fig. 5n**), consistent with the observation in *Wwp2* KO mice^78^. In addition, we observed a similar trend of IL-6 level increase after using another three stimulants, including Pam3CSK4 (a synthetic triacylated lipopeptide and Toll- like receptor 1/2 agonist, **Supplementary Fig. 9b**), Lipopolysaccharide (LPS, an outer membrane component of gram-negative bacteria and a Toll- like receptor 4 agonist, **Supplementary Fig. 9c**), and R848 (a tricyclic organic molecule and an agonist of TLR7/TLR8, **Supplementary Fig. 9d**). However, the increases after stimulations did not reach statistical significance (**Supplementary Fig. 9b-d**).

We studied the evolution of the SV at the *WWP2* locus based on the genotyping results of the high-coverage SRS data of 3,201 samples from 26 populations in the 1000 Genomes Project^45^ (**Fig. 6a and Supplementary Table 4**) and samples from four archaic humans (**Fig. 6b**) as well as the alignment of the human and chimpanzee reference genomes at this locus (**Methods**). We observed that the SV was not present in the genomes of chimpanzees (**Supplementary Fig. 4b**) and the four archaic humans (**Fig. 6b**). In addition, it is rare in African populations (average AF = 0.03) but common (average AF >0.2) in all non-African populations of the 1000 Genomes Project (**Fig. 6a**). Similar to the observation in the *GSDMD,* we identified the signature of positive selection at this SV in multiple non-African populations including the Han based on the genome-wide distribution of *F*ST (**Supplementary Fig. 5; Methods**). Collectively, our results suggest that this SV likely emerged at the common ancestor of modern humans and underwent positive selection after humans migrated out of Africa (**Fig. 6b**).

**Fig. 6:**
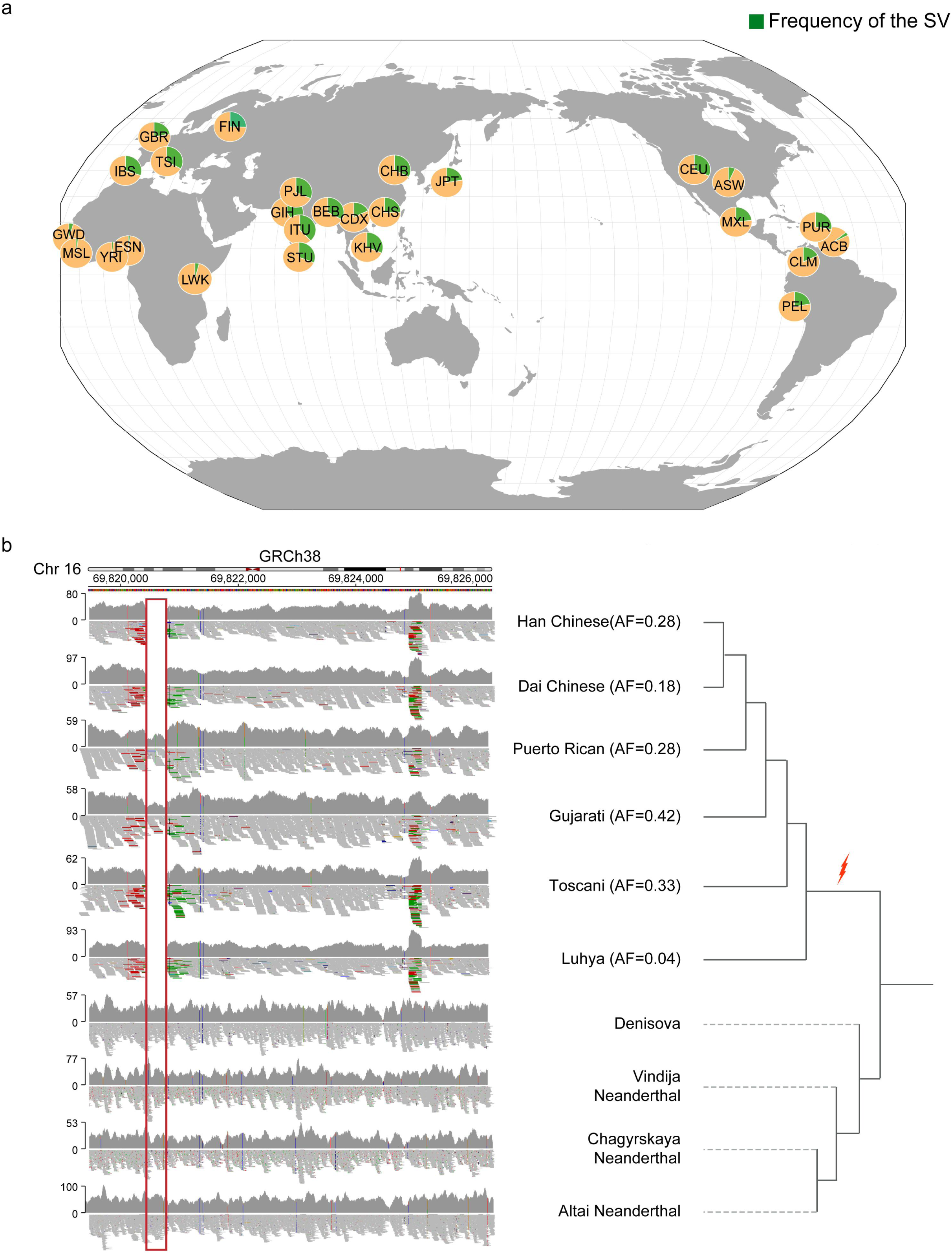
Evolution and demographic distribution of the SV at the *WWP2* locus. **a**, The allele frequency of the SV (indicated in green) in the genomes of 3,201 samples from 26 populations in the 1000 Genomes Project^45^. In the pie charts, the frequencies of the reference and deletion alleles are depicted in yellow and green, respectively. **b,** The read-mapping results around the complex SV at the *WWP2* locus in four archaic human genomes and six representative modern human samples. The Han Chinese sample who carries a homozygous SV is from the present study. We obtained the high-coverage SRS data of one Chinese-Dai individual in Xishuangbanna (CDX, ID: HG01798) who carries a homozygous SV, one Gujarati Indians in Houston, TX (GIH, ID: NA20854) who carries a heterozygous SV, one Toscani in Italy (TSI, ID: NA20585) who carries a homozygous SV, one Puerto Rican in Puerto Rico (PUR, ID: HG01047) who carries a heterozygous SV, and one Luhya in Webuye, Kenya (LWK, ID: NA19318) who carries a homozygous SV. The SRS read-mapping results suggest that the SV (indicated by the red box) is not present in the genomes of the three Neanderthals and the Denisovan. The phylogeny was constructed based on previous studies^2,119–123^. Due to its presence in the genomes of all modern human populations and its absence from the four archaic human genomes, this SV is likely to have emerged in the common ancestor of modern humans (red arrow).

## Discussion

In this study, we investigated the genome-wide SV diversity of 945 samples of Han ancestry using LRS. Our large-scale LRS-based SV detection approach revealed tens of thousands of unreported SVs, many of which were predicted to be functional and of potential biomedical relevance. Moreover, we likely cataloged a majority of common SVs in the Han population. The SV catalog from this cohort can thus help prioritize variants identified in clinical sequencing and serve as a reference panel for incorporating SVs into future GWAS^12^. Furthermore, by integrating these Han Chinese SVs with additional diverse populations, we can gain new insights into human evolution and local adaptation from an understudied SV perspective, complementing existing knowledge primarily based on SNPs. Therefore, the comprehensive SV dataset compiled here represents a valuable genomic resource with diverse applications across life sciences and medicine research.

Our study provides insights into the complex mosaic structure of genomic variation in the Han (**Fig. 2**). Tracing the ancestral origins of SVs reveals contributions spanning highly divergent evolutionary timescales, from millions of years in our hominin ancestry to ongoing mutation processes. Analyses of the functional effects and phenotypic consequences of variants emerging at different periods will shed further light on the genetic changes involved in the origin and diversification of anatomically modern human populations^79^.

Aligning with the findings in previous studies^3,22–26^, our analyses suggest that a substantial portion of phenotypic diversity and differences in disease susceptibility among humans can be attributed to SV-mediated genomic alterations. In addition, we identified the causal SVs that influence *WWP2* and *GSDMD* and their associated phenotypes in humans. Pinpointing causal variants for complex traits has proven remarkably challenging, mainly because the causal variants normally reside in many closely correlated variants due to linkage disequilibrium. Yet doing so provides pivotal insights into key questions surrounding modern human evolution, such as the origin and evolutionary trajectory of a phenotype and the genetic basis of disease susceptibility disparity in modern humans. Despite their prevalence and functional importance, SVs have not been widely used in current GWAS, which remains a primary method for studying complex traits/diseases but relies primarily on SNPs^12^. This has further increased the difficulty of identifying causal variants underlying complex traits or diseases. For example, while our results indicate that the SV in *WWP2* is a novel locus associated with craniofacial polymorphism in humans (**Fig. 5k**), searches of the EBI GWAS Catalog through November 2023 have not implicated any variants in *WWP2* in previous genome-wide association studies of human craniofacial variation. Therefore, there is a strong need to integrate SVs into future GWAS when studying complex traits in humans^12^.

We also identified the phenotypes reported in previous KO mouse models can be misleading or not replicable in humans and humanized mice. For example, prior mouse models exhibited decreased bone density with *Gsdmd* deficiency^55^. However, we revealed the opposite effect in human and mouse carriers of the SV in *Gsdmd*, who displayed significantly increased bone mineral density compared to non-carriers. This disconnect was further highlighted with *Wwp2*. We showed that both human and mouse carriers of the *Wwp2* structural variant have reduced height and weight, yet the abnormal craniofacial and dental changes reported in *Wwp2* KO mice^66^ were not observed in human carriers or our humanized mouse model. One possibility to explain this discrepancy could be caused by a compensatory effect in which knocking out *Gsdmd* or *WWP2* may activate the expression of other members of the gene family. This compensation may not occur or may be less pronounced when knocking out the enhancer of the gene, where some level of expression remains. However, these results could also be explained by knockout experiments, which often remove a large genomic segment and can generate confounding phenotypes by simultaneously disrupting other genes directly or the regulatory elements of other genes^68^. These discrepancies emphasize the need to validate model organism phenotypes in human genetic data.

Our findings suggest a dynamic interplay between genetic variants and environmental pressures that, wherein genetic variants that were adaptive in the past can become maladaptive in modern environments^80^. For example, while emerging at different times, the causal SVs in the *WWP2* and *GSDMD* genes are both rare in Africans while showing signs of positive natural selection in numerous non-African populations (**Figs 4 and 6**). The *WWP2* variant yields increased inflammatory responses and adiposity for its carriers, particularly the accumulation of visceral fat. This may have provided an evolutionary advantage during human migration and adaptation to new environments that often lacked reliable food sources or posed novel pathogenic threats. However, increased visceral fat is associated with an increased risk for multiple diseases in modern societies, such as high blood pressure, obesity, elevated cholesterol, and insulin resistance^81^. The SV in the *GSDMD* locus is associated with increased bone density. Denser, stronger bones would have reduced susceptibility to fractures and improved load- bearing capacity, which could be beneficial in physically demanding environments or situations where physical prowess is necessary for survival, such as hunting, combat, or escaping predators. However, osteosclerotic bones also tend to be misshapen and abnormally enlarged. This progressive skeletal deformity can cause reduced mobility, balance impairments, and difficulty with fine motor tasks^82^. Overall, by unraveling the phenotypic effects and evolutionary origins of variants like the SVs in *WWP2* and *GSDMD*, we gain novel insights into genomic diversity and local adaptation during human migration across the globe. Reconstructing this complex history is also key to understanding geographic differences in disease and designing personalized medical strategies tailored to patients’ ancestry.

Our findings provide a rapid and cost-effective predictive biomarker that could enable personalized risk stratification when treating cancer before cisplatin administration and other *GSDMD*-mediated pyroptosis in organ injuries. Using a PCR-based screening for the SV affecting a conserved enhancer regulating the expression of *GSDMD* in patients, clinicians may be able to adjust cisplatin dosing or provide alternative regimens for susceptible patients and thus reduce the risks of acute kidney injury. As cisplatin is a widely used and effective chemotherapeutic against multiple solid tumor types, mitigating its toxicity to the kidney and other organs could significantly improve clinical outcomes and quality of life for many patients. In addition, the dysregulation of *GSDMD* has also been implicated in multiple organ injuries^83^, such as liver^84,85^, brain^86,87^, spinal cord^88–91^, heart, and vessel^83,92–94^, the SV can serve as a potential novel intervention target by selectively controlling *GSDMD* expression in relevant cell types and thus allowing interventions before downstream injury processes propagate^95–97^.

The present study has some limitations. First, the functional divergence between human and mouse orthologous genes may have misled us to interpret the functional impacts of some SVs. Experiments utilizing conditional knockout or knocking in the human variants in mouse models ^98^ or knockdown of genes in human organoid models using siRNA or shRNA, or gene editing of human embryonic stem cells or induced pluripotent stem cells using CRISPR/Cas9 can overcome some limitations of conventional knockouts and further elucidate gene functions in a human context. Additionally, our estimates of SV emergence and evolution are constrained by factors including the availability of only four sequenced archaic human genomes and genetic polymorphisms not represented within the 1000 Genomes Project samples. Moreover, since we primarily focused on studying the common SVs, we may have missed some rare, but functionally important SVs. As we enter the ’phenomics’ era^99,100^, characterized by the acquisition of high-dimensional phenotypic data in diverse global populations using various ’- omics’ datasets and the ongoing advancements in both *in vitro* and *in vivo* technologies as well as genomic data from underrepresented populations, we anticipate that the functions of SVs will be elucidated in a broader range of human populations in the future^80^.

## Methods

### Inclusion and Ethics

The study complied with all relevant regulations for working with human subjects in China. The Ethics Committee of the School of Life Sciences, Fudan University, Shanghai, China approved the study. Participants were recruited to a project studying physical anthropology diversity in China funded by the Ministry of Science and Technology of the People’s Republic of China (2015FY111700). Informed consents were approved by all participants.

### Sequencing library preparation

We extracted DNA from whole blood using a TGuild Blood sample genomic DNA kit from TIANGEN®. The ONT libraries were prepared as follows: 2‒5 µg genomic DNA was sheared to ∼20‒30 kb fragments in a g-TUBE (#520079, Covaris) by spinning twice for 2 min at 7,000 rpm in an Eppendorf MiniSpin centrifuge. The samples were mixed with 6.5 μl of NEBNext FFPE DNA Repair Buffer, 2 μl of NEBNext FFPE DNA Repair Mix (New England Biolab, M6630), and 3.5 μl of nuclease-free water (NFW) and incubated at 20 °C for 15 min for DNA repair. We re- pooled and cleaned up the samples using a 0.8× volume of AMPure XP beads (Beckman Coulter) according to the manufacturer’s instructions. The purified-ligated DNA was resuspended in 15.5 μl ELB (SQK-LSK108). A 1-μl aliquot was quantified by fluorometry (Qubit) to ensure ≥ 500 ng DNA was retained. The final library was prepared by mixing 35.0 μl RBF (SQK-LSK108), 25.5 μl LBB (SQK-LSK108), and 14.5 μl purified-ligated DNA.

### ONT sequencing and base calling

We conducted long-read sequencing of 945 Han Chinese individuals using an ONT PromethION sequencer with a 1D flow cell and protein pore R9.4 1D chemistry according to the manufacturer’s instructions. Base calling was performed using Guppy version 3.2.0^101^ with the “flipflop” algorithm on the PromethION compute device.

We measured the head length of the samples using a spreading caliper.

### Whole genome sequencing, phenotypic and immunogenic and metabolic profilings of an independent cohort

We conducted whole genome sequencing of 1,016 samples using the BGI-DNBSEQ-T7 platform. Briefly, the DNA samples were extracted from whole blood. Paired-end libraries (with insert size around 280 bps) were prepared based on the manufacturer’s instructions. For each sample, we sequenced 100 bps at each end and obtained >90 Gb sequencing reads, corresponding to 30X coverage. We removed the reads <25 bps, or ≥10% unidentified nucleotides (N), or 50% bases having phred quality <5, or >10 nt aligned to the adapter/primer/barcode/index, allowing ≤10% mismatches, or show significant biased sequence composition difference such as the reads between the A/T and G/C ratio greater than 20%, or potential PCR duplicates.

The sequencing reads were mapped to the human reference genome GRCh38 using BWA-MEM mode (version 0.7.17)^102^ with default parameters. The mapping results were processed using SAMtools version 1.15.1^103^ to sort and index. Finally, we used GATK version 4.1.7.0^104^ to mark duplicated reads. We genotyped the LRS-based SVs in this cohort using Paragraph version 2.3^46^ with default parameters.

We also measured the standing height of the samples in this cohort. Whole-body BMD of 630 volunteers was assessed by dual-energy X-ray absorptiometry (DXA). The BMD (g/cm^2^) measurement was performed by the Lunar iDXA (GE Healthcare, General Electric Co., Boston, MA, USA) using standard testing procedures.

Inflammatory factor levels were measured as described in a previous study^105^. Specifically, venous blood was collected from healthy individuals into sodium heparin tubes for whole blood stimulation assays. Blood was diluted 5-fold in a 48-well plate pre-filled with RPMI media (10% FBS, 1% penicillin/streptomycin) and four stimulants: Pam3CSK4 (100 ng/mL), flagellin (100 ng/mL), LPS (10 ng/mL), R848 (500 ng/mL), or no stimulant (negative control). Plates were incubated at 37°C with 5% CO2 for 24 hours. Supernatants were collected by centrifuging at 500 g for 10 minutes and immediately frozen at -80 °C until analysis. The levels of IL-6 were quantified using ELISA (Biolegend #555220) per manufacturer protocol.

We purchased PG (15:0/15:0), PI (17:0/14:1) from Sigma-Aldrich (St. Louis, MO, USA) and the LipidyzerTM kit from (SCIEX, Chromos, Singapore). Sinopharm Chemical Reagent Co., Ltd. (Shanghai, China) provided the HPLC grade Chloroform. Merck (Darmstadt, Germany) supplied Methanol (MeOH), isopropanol (IPA), methyl tert- butyl ether (MTBE) and dichloromethane (DCM). Sigma-Aldrich (St. Louis, MO, USA) obtained acetonitrile (ACN), formic acid (FA), ammonium hydroxide (NH4OH) and ammonium acetate (NH4OAc). We purified ultra-pure water using the Milli-Q water purification system (Millipore, MA, USA).

We extracted 8 uL plasma samples and 8 uL lipid standard mixture using the reported Matyash method with MTBE/methanol/water (10:3:2.5, V/V/V)106. We analyzed all lipids based on previously reported methods^107,108^ with some minor modifications. Here we used an LC-MS-based method on a QTRAP 6500 plus mass spectrometer (Sciex, USA) coupled with an ultrahigh performance liquid chromatography system LC30A (Shimazu, Japan). We employed a BEH HILIC column (2.1 100 mm, 1.7 μm) and Kinetex C18 column (2.1 100 mm, 2.6 μm), where appropriate, with solvent flow rates of 0.5 mL min-1 and 0.3 mL min-1, respectively. Mobile phases included H2O/ACN (5:95, V/V) and H2O/ACN (1:1, V/V), both containing 10 mM NH4AC for the former, while H2O/MeOH/ACN (1:1:1, V/V/V) containing 7 mM NH4OH and IPA containing 7 mM NH4AC for the latter. We used a 12-min gradient elution for both columns. The samples injection volumes were 2 μL and 1μL respectively, and we set the column temperature at 45°C.

We set the MS parameters of MRM acquisition mode for lipid species as previously described: collision gas (CAD), medium level; CUR, 40 psi; the GS1 and GS2, both 55 psi; the ion spray voltage value was relatively set at 5500 V or –4500 V; Turbo V source temperature, 350 °C; DP, ± 70 V. We optimized the collision energy values for these subclasses, respectively.

We used Analyst (v1.7, SCIEX, Chromos, Singapore) and OS (v1.7, SCIEX, Chromos, Singapore) to quantify the data for each lipid species. OS extracted the peak area and tR from the raw data. We calculated the lipid concentration by dividing the peak area of each lipid species by the peak area of the corresponding internal standard (IS) and then multiplying by the concentration value of the IS.

### LRS read mapping

The sequencing reads were mapped to the human reference genome (GRCh38.p13, without alternate sequences) using NGMLR version 0.2.7^28^ with parameters specifically tailored for ONT reads. The mapping reads were sorted and indexed using SAMtools version 1.17^103^.

### LRS-based SV calling and quality control

To account for the moderate coverage of our samples, a joint-calling strategy was used to call SVs in this study. Specifically, we first conducted SV calling per sample using cuteSV version 1.0.13^29^ with parameters recommended for ONT reads by the developers. We merged SVs whose breakpoints were within 500 bps across individuals using SURVIVOR version 1.0.6^30^. Based on the merged SV set, we used LRcaller version 1.0^23^ to re- genotype each sample and then remerged all SVs using the BCFtools (version 1.17) merge command^103^ to generate a complete SV set. To obtain high-quality SV calls, we removed SVs located in centromeric, pericentromeric and gap regions in GRCh38.

### SV genotyping using SRS data of the 1000 Genomes Project and four archaic humans

We extracted the SRS reads around the *GSDMD* (chr8:143,541,891‒143,564,066, GRCh38) and *WWP2* (chr16:69,810,428‒69,834,903, GRCh38) loci from the high-coverage BAM files of the samples of the 1000 Genomes Project from https://ftp-trace.ncbi.nlm.nih.gov/1000genomes/ftp^45^ using SAMtools version 1.15.1^103^. We conducted SV genotyping using Paragraph version 2.3^46^ with default parameters.

The BAM files of four archaic humans were downloaded from http://ftp.eva.mpg.de/neandertal/^61,62,106^ and http://cdna.eva.mpg.de/denisova/^64^. As the sequence read data for the four archaic humans were mapped to the GRCh37 human reference genome, we converted the breakpoint coordinates identified in this study from GRCh38 to GRCh37 using the liftOver tool in the UCSC Genome Browser^77^ for comparative analyses.

To examine whether SVs that are fixed (AF = 1) or nearly fixed (AF >0.9) in the Han population also exist with high frequency in other global populations. We genotyped all the high-frequency SVs in the Han population using Paragraph version 2.3 with default parameters^46^ based on 2,503 samples in the 1000 Genomes Project^45^ and 50 samples were randomly chosen from an independent cohort in our present study. The allele frequency of each SV was calculated using BCFtools (version 1.17)^103^ with the command of “bcftools +fill-tags sv.vcf.gz -- -t AF”.

To explore the origin of SVs in the Han, we genotyped 110,863 SVs (after excluding 372 TRAs and 53 SVs containing non-ATGC bases in their alternative sequence that cannot be genotyped by Paragraph^46^) using Paragraph version 2.3 with default parameters^46^ based on 2,503 unrelated high-coverage sequencing genomes of the 1000 Genomes Project^45^ as well as four archaic humans^61,62,64,106^.

### A comparison to the SVs in the chimpanzee genome

We downloaded the VCF file named ’AG18359_ONT.hg38.vcf’ from a previous study^47^, in which the authors mapped the ∼30X ONT reads of chimpanzee AG18359 lymphoblastoid cell lines to the human reference genome GRCh38 using minimap2^107^ (v2.17-r941) and identified 95,245 SVs (length ≥50 bps) using Sniffles^28^ (v1.0.11). After converting the VCF to BED format, we intersected these SVs with those identified in the chimpanzee genome using BEDTools (v2.30.0)^108^ applying a 50% reciprocal overlap threshold. This analysis revealed 9,508 SVs of the same type shared between the chimpanzee genome and our cohort.

### Estimations of false discovery rate (FDR) and false negative rate (FNR)

We successfully validated 91 SVs, including 39 insertions, 42 deletions, four inversions, and six duplications, with sizes ranging from 53 to 81,592 bps using a PCR-based approach. To estimate the boundaries of the FNR and FDR of our dataset, we adopted a method from Beyter et al., 2021^23^, which is based on comparisons with the high-frequency (AF >0.5) SVs in the global populations. Specifically, we used two datasets (Audano et al., 2019^1^ and Human Genome Structural Variation Consortium Phase 2 (HGSVC2)^3^, which respectively characterized SV diversity of 15 and 35 individuals of diverse ancestry, into the comparisons. We used GORpipe version 3.10.1^109^ with the parameters “A.gor | join -snpseg -f 500 B.gor”, which examines a variant in dataset A that is also present (within 500 bps of the start position of the variant discovered in dataset A) in dataset B.

### Identification of the unreported SVs in the present study

We examined that whether the SVs in the present study were reported in five publicly available SV datasets, including gnomAD^31^, Audano et al., 2019^1^, HGSVC2^3^, Wu et al.^22^, and dbVar^33^ (as of May 19, 2023). We used liftOver in the UCSC Genome Browser^77^ to convert the coordinates of SVs identified by gnomAD from GRCh37 to GRCh38. When a reciprocal overlapping rate between an SV in our and published datasets ≥50%, we defined it as reported before.

The functional impacts of these unreported SVs were predicted based on gene coordinates of the RefSeq databases (as of August 17, 2020)^110^ using AnnotSV (version 3.1.1)^48–50^. We obtained the intersections of unreported SVs with transcription factor binding site, and the chromatin state segmentations for cell lines GM12878, H1-hESC, HepG2, HMEC, HSMM, HUVEC, K562, NHEK, and NHLF from ENCODE project^34^ after using liftOver^77^ software to convert the coordinates of SVs from the GRCh38 version to GRCh37 based on ANNOVAR (version 2019Oct24)^35^.

### Analyses of SV breakpoints and repeat elements

We downloaded the repeat elements annotation file of GRCh38 from the UCSC website^77^ (https://hgdownload.soe.ucsc.edu/goldenPath/hg38/database/rmsk.txt.gz). The intersect module in BEDTools version 2.30.0^108^ was used to examine whether the SV breakpoints overlapped repeat elements or not.

### Calculation of the distances between SVs and telomere regions

The telomere positions on each chromosome were downloaded from the UCSC online website^77^ using the Table Browser tool^111^ (https://genome.ucsc.edu/cgi-bin/hgTables). The distance between SVs and telomeres was defined as the minimum distance between SV breakpoints and telomeres on both sides.

### A gene-based function annotation and gene ontology enrichment analysis

A gene-based annotation was performed using AnnotSV (version 3.1.1)^48–50^ with the gene coordinates of the RefSeq databases (as of August 17, 2020)^110^. We performed gene ontology and Kyoto Encyclopedia of Genes and Genomes (KEGG) pathway enrichment analysis using the 2023 version of the DAVID web server^51,52^ (https://david.ncifcrf.gov/). The gene ontology and KEGG pathways with false discovery rate (FDR) <0.1 were considered as significant.

### Visualization of epigenomic data for SVs at *GSDMD* and *WWP2* loci

We used UCSC Genome Browser (https://genome.ucsc.edu/)^77^ to visualize epigenomic data for SVs at the *GSDMD* and *WWP2* loci. We obtained the enhancer regulatory elements annotated by GeneHancer database^53^, ChIP-seq signals of H3K4me1, H3K27ac, CTCF, and transcription factor binding sites from the Roadmap Epigenomics project^73^ and ENCODE project^34^, regulatory elements from ORegAnno^112^, and Hi-C chromatin interactions reported by Rao et al.^113^. To visualize the Hi-C interactions, we first used the Table Bowser tool (https://genome.ucsc.edu/cgi-bin/hgTables)^111^ in the UCSC genome browser to download all the Hi-C interactions in the regions (*WWP2* locus: chr16:69,685,000‒70,025,000, *GSDMD* locus: chr8:144,620,000‒144,655,000, GRCh37).

We then used the intersect module of BEDTools version 2.30.0^108^ to obtain the Hi-C interactions between the SV region and the regions located within 1 kb of the transcription start sites of *WWP2* and *GSDMD*. We uploaded the remaining interactions to the UCSC Genome Browser website (https://genome.ucsc.edu/)^77^ for visualization.

### Identification of the common SVs located in the annotated enhancer regions in the human reference genome

Using the intersect module of BEDTools toolkit version 2.30.0^108^, we searched the common SVs (with MAF ≥5%) in our dataset that completely overlapped the enhancer annotations from ENCODE project^34^, Roadmap Epigenomics project^73^, and GeneHancer database^53^.

### Exploring the functional significance of genes regulated by common SVs in mice

We explored the potential functions of common SV-regulated genes based on the Mouse Genome Informatics database^65^. We first mapped the genes regulated by SVs to homologous genes in mice based on the HMD_HumanPhenotype.rpt file downloaded from https://www.informatics.jax.org/downloads/reports/index.html#pheno. After excluding genes that were lethal or genes that had no obvious phenotypic effects in mouse KO models, we then classified the phenotypic changes in the remaining genes based on the MGI_GenePheno.rpt file downloaded from https://www.informatics.jax.org/downloads/reports/index.html#pheno.

### Local assembly of the SV at the *WWP2* locus

We extracted LRS reads spanning over the breakpoints from the bam files of the carriers of the SV in our cohort, and used Flye version 2.9.2^114^ to perform local assembly. The assembled contig was aligned to the reference genome using the online BLASTN (https://blast.ncbi.nlm.nih.gov/Blast.cgi) based on which the dotplot in Fig. 5d was generated.

### A SINE fusion event in *WWP2* causes false positive detection of 4.5 kb deletion

We utilized Manta version 1.6.0^115^ and Delly version 1.1.8^116^ to detect SV around the *WWP2* locus based on high-coverage BAM files of the samples of the 1000 Genomes Project from https://ftp-trace.ncbi.nlm.nih.gov/1000genomes/ftp^45^. In total, 138 samples were used (sample IDs are listed in **Supplementary Table 8**). We set the “--generateEvidenceBam” parameter for Manta and “-d” parameter for Delly to report the reads that support the detection of the SV. Both Manta and Delly reported a ∼4.5 kb deletion (Manta: chr16:69,820,428‒69,824,903, Delly: chr16:69,820,426‒69,824,899). We found that the reads that support this deletion originated from the fused SINE element region using Integrative Genomics Viewer version 2.11.1^117^. We also observed a ∼4.5 kb deletion was reported by cuteSV in our cohort.

### Alignments of the human and chimpanzee reference genomes around the SVs at the *WWP2* and *GSDMD* loci

We obtained homologous regions between the reference genomes of human (GRCh38) and chimpanzee (PanTro6, January 2018 assembly) at the *WWP2* and *GSDMD* loci using Lift Genome Annotations^111^. In addition to the sequence within each SV, we also extracted 2000 bps upstream and downstream of the SV breakpoints. We then aligned the sequences using the NCBI online BLASTN tool (https://blast.ncbi.nlm.nih.gov/Blast.cgi).

### Positive selection scan of the SVs in global populations

We downloaded the SV calls in a previous study of SV diversity in global population using LRS data^3^ from https://ftp.1000genomes.ebi.ac.uk/vol1/ftp/data_collections/HGSVC2/release/v2.0/integrated_callset/variants_freeze4_sv_insdel_alt.vcf.gz. We extracted a total of 102,118 insertions and deletions located in autosomes using BCFtools (version 1.17)^103^ view command. We merged the insertion call at the *WWP2* locus with this dataset, since the deletion at the *GSDMD* locus is reported.

To test whether the SVs at *WWP2* and *GSDMD* were under positive selection in global populations or not, we randomly selected 50 samples per population in the 1000 Genomes Project and downloaded their high- coverage bam files from https://ftp-trace.ncbi.nlm.nih.gov/1000genomes/ftp^45^ (**Supplementary Table 7**). Additionally, 50 samples were randomly chosen from the independent cohort in our present study. We genotyped all the SVs in the merged dataset using Paragraph version 2.3 with default parameters^46^. The *F*ST were calculated between the African populations and each of the non-African populations using VCFtools version 0.1.16^118^ with the following parameters “--weir-fst-pop African_population.txt --weir-fst-pop non_African_population.txt”. We then analyzed the percentile distribution of *F*ST for SVs at *WWP2* and *GSDMD* by population.

### PCR and Sanger sequencing of the SVs at the *WWP2* and *GSDMD* loci

One sample carrying heterozygous SV at *WWP2* locus and one sample carrying heterozygous deletion at *GSDMD* locus were used to validate the SVs at the *WWP2* and *GSDMD* loci. PCR primers were designed outside of the SV region. Therefore, we can get wild-type and SV-affected sequences for the downstream luciferase reporter assays. Specifically, primers with homology arms of pGL3-Promoter vector were designed using Primer- BLAST (https://www.ncbi.nlm.nih.gov/tools/primer-blast/index.cgi) and were synthesized by TsingKe Biotech Corp (Shanghai, China). The homology arms were designed to have sequence complementarity with regions adjacent to the XhoI restriction site on both the target vector and the DNA fragment to be inserted. The empty pGL3-Promoter vector was kindly provided by Dr. Xiaoyang Zhang at Fudan University. Considering the sequences around the *WWP2* locus are highly repetitive and contain SINE elements, the target region was amplified using touchdown PCR to improve amplification efficiency and reduce non-specific amplification. Specifically, the PCR amplification cycling consisted of an initial denaturation step at 95°C for 3 minutes, followed by 30 cycles of the following steps: denaturation at 95°C for 15 seconds, annealing at temperatures ranging from 62°C to 55°C for 15 seconds, decreasing the annealing temperature by 1°C per cycle for the first few cycles, and finally, extension at 72°C for x minutes (1-2 kb/min). 2* Phanta Max Master Mix (Dye Plus) (Vazyme, P525-01) was used, with 50 ng of genomic DNA (Homo sapiens) as the template in a final reaction volume of 50 μl. All PCR products were subjected to agarose gel electrophoresis. Trans2K^®^ Plus II DNA Marker (Trans, BM111-01) was used as molecular weight standards for bands of PCR products.

We performed Sanger sequencing based on the PCR products at the *WWP2* and *GSDMD* loci. The sequence at the *WWP2* locus is:

ATGGGTAGCGACGGCAGTTGATCTGAACTCAGGATCACTCGTGTTACACTGCAATCGCGTGTCGCCCTTTCAGAGACCGCTAAAGACAGCCTGAAATCCCAAAGCCTGGCATCTGGTGCCAATCAAAAAAACAGTAGTCGCTGATGTAACTGCAACTCGATCAGGGCAAAATGAAACAACGGCTTGCATTTGGCGGCTGGAACTGGGCTCAGCCCATAGCCTCGCCGGAGTGGGCAGCACGCACCACACTGGTCCTGCTGTCAGCCTCCGAGGTGAGCCGGACACTGGCTGCAAAGACCTAAGTCACAGGAAAAGACGTTCATCCTCAGTAATTAGAGGAGAAAGTCACTCTTTCAACGTCGTGTGTGTGTGTGTGTGTGTGTGTGTGTATATGCATGTATTCATGGTTTTGTGTTGTAGGTACACAAAAGTTGTTCACTTTTGTGTATGAAGTCATTGCAAACACAAGCACAAATATACACATAAAAGCTACAGATGCAAGGCCGAGCACAGTGGCTCATGCCTGTAATCCCAGAGCCTGAGGTGGGTGGATCACCTGAGCTCAGGAGTTCGAGACCAGCCTGGCCAACAAAGTGAAACCCCATCTCTACCAAAAATACAAAAATTAGTCAGGCATGGTTGTGCATGCCTGTAATCCCAACTACTAAAAATACAAAAAAATTAGCCAAGCACAGTGGCACATGCCTTTAATCCCAGCTACTCAGGAGGCGGAGGCAGGAGAATCACTTGAACCCAGGAGGGCGGAGGTTGCAGTGAGCTGAGATGCACCACTGCACTCCAGCCTGGGCAACAGAGTGAGACTCCATCTCAAAAAAATAATAATAACAAAAATAAAGGAGACAGGGGTCCCACTCTGTCACCAGGCTGGAGTACAGTGGCATGATCACAGTCACTGGATCTCGACACCCAA (strand: minus)

The sequence at the *GSDMD* locus is:

GGACTTAATGTAGTGGGCCTCTGTGTTCCTGTGTGTGCACATGTCGCTGTGTGTGTTCATGTGCAGGTCACGGTGCGCAGCCGGTGCCACGCCCGTGGCTGTCCGCTGCAGGAAGGGGAGCCCATCTGGGCCTCCTCCCTCCTCCCCTCCTCCCGCTTTTCTCCTTTCTTTTGCAGCAAAATTCCTGGAAAGCCCTGTACACTGCACTCCAGCCTGGGTGACAGAGCAAGAATCCATCTCTAAAATAAAAATATACTTTCTCCCAAGGTCCCCAGCCCAGAGGGTGTGAGGGCCTTGCAGTGGGAGGTAGGTGTCACTGCGCATCCGTGACAGTGGGGAGAGTGGGATGAGGGGGACCCACCAGACCTCTAGAGCAGTTTTCTCCCACTGTCACTTTCTCCCTCCATAAAAGGGG (strand: minus)

### Quantitative real-time reverse transcription (RT-qPCR)

We examined the impacts of the SVs on the expressions of *GSDMD* and *WWP2* based on RNAs of a Chinese quartet family, the parents and two identical twins (http://chinese-quartet.org/)^76^. Total RNAs were extracted from whole blood. cDNAs were reversed-transcribed using a HiScript® III RT SuperMix for qPCR (+gDNA wiper) kit (Vazyme, R323-01), followed by amplification of cDNA using ChamQ Universal SYBR qPCR Master Mix (Vazyme, Q711-02). The relative mRNA levels of genes were quantified using the 2^-ΔΔCt^ method, with normalization to *GAPDH*. Primers are listed in **Supplementary Table 9.**

### Recombinant plasmids construction

DNA fragments were prepared by PCR amplification or synthesized by Tsingke Biotech Corp (Shanghai, China). The recombinant plasmids were constructed by homologous recombination using ClonExpress II One Step Cloning kit (Vazyme, C112) according to the manufacturer’s instructions. The reaction mixtures were then transformed into F-DH5α (WeiDi, WD-DF1001M) using heat shock. Transformed cells were selected on agar plates containing ampicillin. Positive colonies were screened and sequenced in both directions by Sanger sequencing to ensure correctness. Plasmids from the positive colonies were isolated using TIANprep Mini Plasmid Kit (TIANGEN, DP103).

### Cell culture, transient transfections and luciferase reporter assays

The HeLa cell line was obtained from the American Type Culture Collection (ATCC) and cultured in Dulbecco’s modified Eagle’s medium (DMEM) (BasalMedia, D211114), which was supplemented with 10% fetal bovine serum (FBS) (Life-iLab, AC03L055) and 1% penicillin/streptomycin (Life-iLab, AC03L332). HeLa cells were maintained at 37°C with 5% CO2 and were plated on 12-well dishes 24 hours prior to transfection and grown to approximately 50% confluency. The cells were transfected with 1 μg of enhancer plasmids and 0.1 μg of Renilla plasmid as an internal control of transfection efficiency using EZ Trans RNA transfection reagent (Life-iLab, AC04L051) according to the manufacturer’s instructions. Empty pGL3-Promoter vector was co-transfected with Renilla as control. Cells were harvested 48 hours after transfection. The cell pellet was fully lysed in reporter lysis buffer (Beyotime, RG027) and the supernatant was acquired after centrifuging at 12,000 rpm for 5 minutes. The luciferase activity was measured with the dual luciferase reporter gene assay kit (Beyotime, RG027) using Synergy™ 2 Multi-Mode Microplate Reader (BioTek). Each assay was performed in triplicate. The relative luciferase activity was determined by dividing the Relative Light Units (RLU) value of Firefly luciferase by the RLU value of Renilla luciferase based on the manufacturer’s prescribed method.

### Mice experiments

Mice (strain: C57/Bl6) were raised and maintained in a barrier facility. All animal experiments were reviewed and approved by the Institutional Animal Care and Use Committee of the East China Normal University, and conducted according to institutional guidelines. Corresponding littermates were used as WT controls in all of the experiments performed with KO mice. We identified the homologous regions in the mice genome using the liftOver tool in the UCSC Genome Browser website^77^. *Wwp2* and *Gsdmd* mutant mice were generated by co-injection of Cas9 mRNA (100 ng/μl; ThermoFisher, A29378) and sgRNA (50 ng/μl) at the Mouse Core of East China Normal University. sgRNAs were generated using the Guide-it™ sgRNA In Vitro Transcription Kit (Takara #632635), with their sequences listed in **Supplementary Table 9**. Genomic DNA was extracted from newborn F0 mice’s toes or tails one week after birth for sequencing, with genotyping primers listed in **Supplementary Table 9**. For the cisplatin-induced injury model, 8-week-old gender-matched wild-type and *Gsdmd* enhancer homozygous KO mice received intraperitoneal one-time injections of cisplatin (25 mg/kg) and were euthanized three days later.

### microCT analysis

For micro-CT analysis, the proximal femur and skull of 7-week-old mice were scanned using a Micro-CT scanner (Skyscan 1272, USA Bruker) with an X-ray energy of 450 μA/50 kV and an isometric resolution of 9 μm. The trabecular bones were assessed for bone mineral density, BV/TV ratio, trabecular number, trabecular thickness, and trabecular separation/spacing using the CT Evaluation Program.

### Histopathology analysis

Kidneys were harvested from mice, rinsed in PBS, fixed in 10% formalin, and then embedded in paraffin. Paraffin-embedded sections were stained with hematoxylin and eosin (H&E) and TUNEL to analyze the histology of samples.

### Statistical Analysis

Statistical analyses were performed using GraphPad Prism software version 8.0.2 (GraphPad Software Inc., La Jolla, CA) and R version 4.1.0. *P* values less than 0.05 were considered statistically significant.

## Data Availability

All data produced in the present study are available upon reasonable request to the authors

## Acknowledgements

The authors thank Prof. Sarah A. Tishkoff at the University of Pennsylvania, Prof. Feng Zhang, Prof. Jin Li, and Prof. Wei Yu at Fudan University for their insightful comments on the manuscript. The authors thank Nextomics Biosciences for covering the sequencing cost of 100 samples. This work was supported by the National Natural Science Foundation of China (grants No. 31970563 and 32370686 to S.F., 32271186 to J.T., 32030020 to S.X., and 82100773 to Y.G.) and National Key Research and Development Program of China (2020YFE0201600 and 2021YFC2500202 to S.F. and 2022YFC2505400 and 2022YFC3400203 to Y.G.), the 111 Project (B13016 to S.F. and L.J.). Shanghai Municipal Science and Technology (grant No. 2017SHZDZX01) to L.J., Shanghai Rising-Star Program (22QA1405900 to Y.G.). F.J.S. is supported by NIH grants (1U01HG011758-01, U01 AG058589). This work was also supported by the Human Phenome Data Center at Fudan University.

## Author contributions

S.F. and L.J. conceived the experiments. S.F., L.J., and F.J.S. designed the experiments. S.F. and Y.G. wrote and revised the manuscript with the help of other authors. J.G. conducted LRS mapping, SV calling, and annotation. H.S. conducted the cellular experiments. Y.G. and K.W. conducted the mice experiments. J.T., Y.M., J.L., and S.W. collected the samples. H.S. and Y.Z. conducted PCR and Sanger validation. H.Q. conducted the head length analysis. Y.G. performed SRS read mapping. J.Z. and F.Q. performed immunological experiments. Q.C., Q.W., and H.T. conducted metabolomics data analysis. J.L. coordinated the collection of the phenotypic data of the independent cohort. All authors read and approved the manuscript.

## Data and materials availability

The data were stored at the National Genomics Data Center, China (https://ngdc.cncb.ac.cn, accession number HRA005917). The VCF file containing all SV calls from this study will be made publicly available upon acceptance of the manuscript for publication.

## Additional Information

Supplementary Information is available for this paper.

Correspondence and requests for materials should be addressed to.

