## Supplementary Figures for "Long-read sequencing of 945 Han individuals identifies novel structural variants associated with phenotypic diversity and disease susceptibility"

**Supplementary Information**

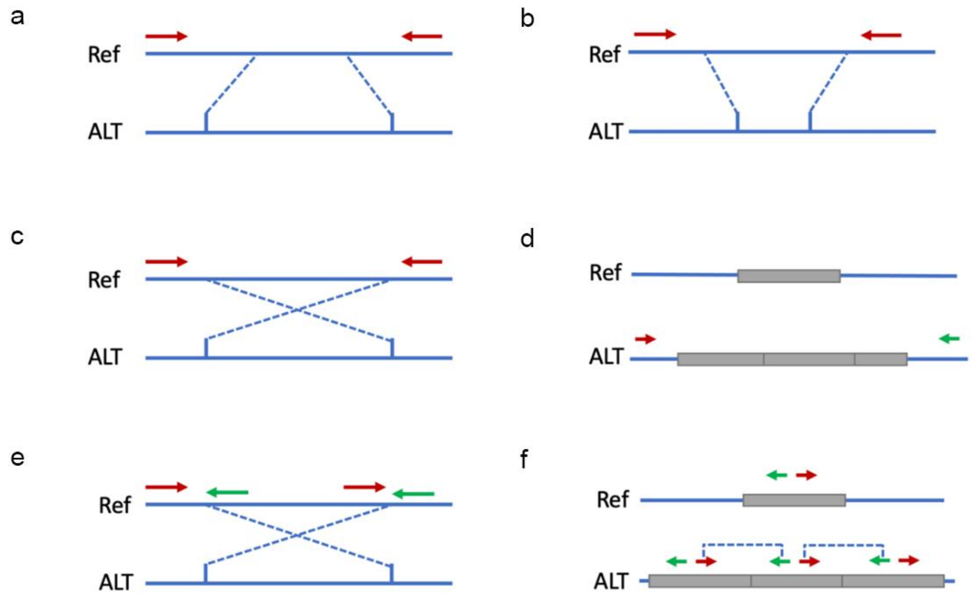


**Fig. S1:** The scheme of structural variants validation using a PCR-based approach. Primers were designed in the regions outside of breakpoints for insertion (**a**), deletion (**b**), inversion (**c**), and duplication (**d**) calls. In addition, we also used both forward or both reverse primers for inversions (**e**) and tail-to-tail primers for duplications (**f**).

Deletion M 1 2 3 4 5 6 7 8 9 10 11 12 13 14 15 16 17 18 19 20 21 22 23

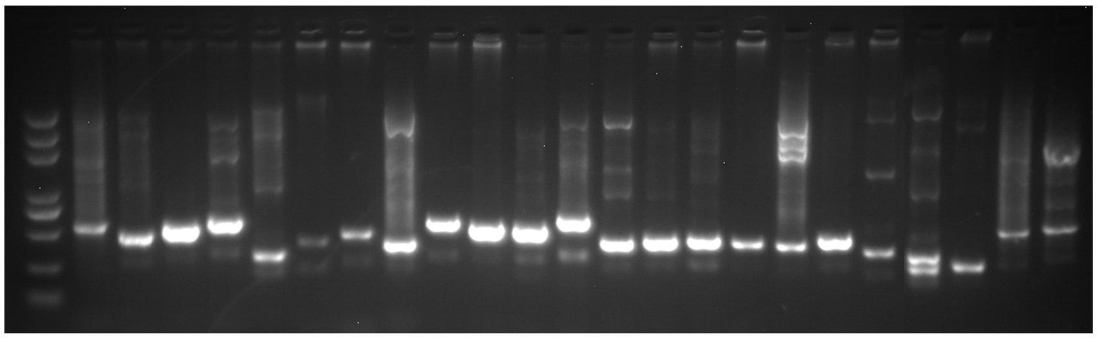


Deletion M 24 25 26 27 28 29 30 31 32 33 34 35 36 37 38 39 40 41 42

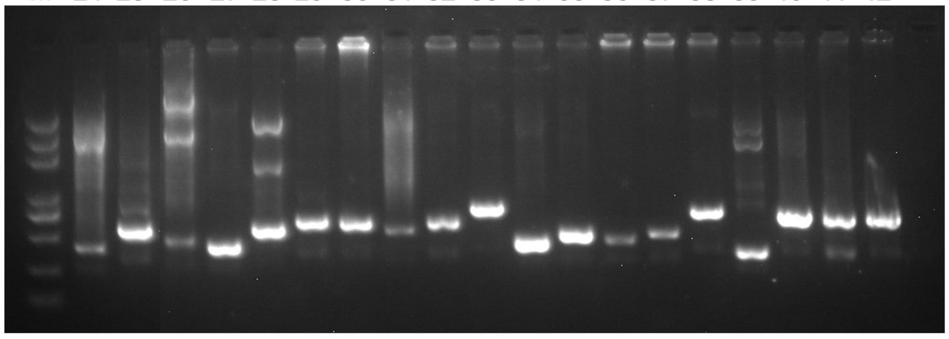


Insertion M 1 2 3 4 5 6 7 8 9 10 11 12 13 14 15 16 17 18 19 20 21 22 23 24

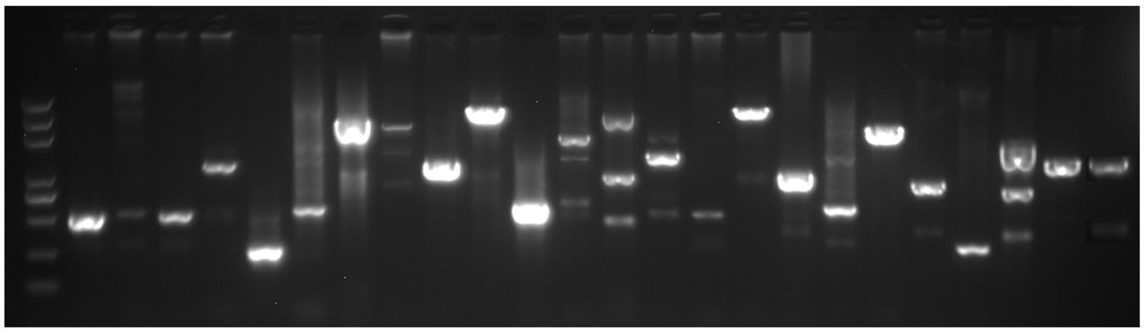


Insertion M 25 26 27 28 29 30 31 32 33 34 35 36 37 38 39

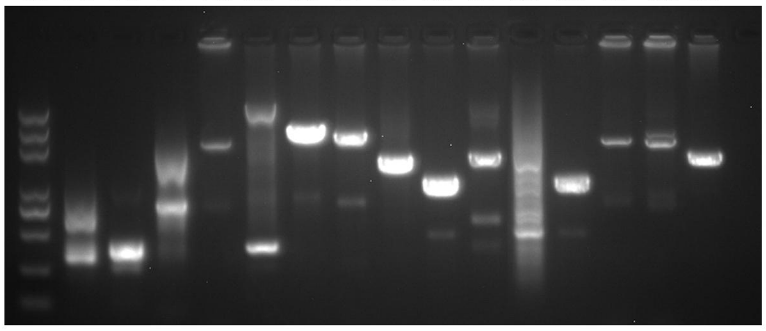

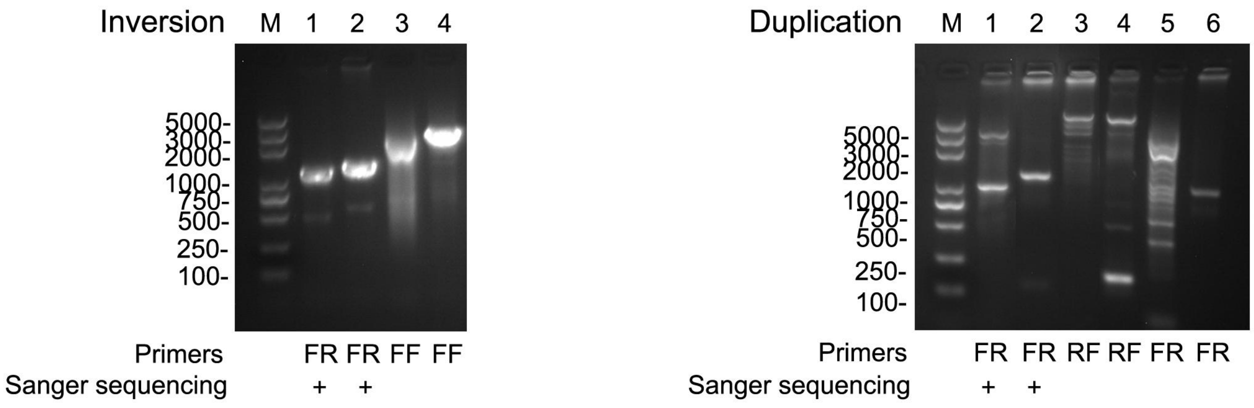


**Fig. S2:** Agarose gel electrophoresis (1.3%) of PCR products of 91 SVs. We verified a total of 91 SVs including 42 deletions, 39 insertions, 4 inversions, and 6 duplications. "+" indicates the PCR product was sequenced using Sanger sequencing. M: Size marker. Besides the forward-reverse (FR) primers, we also used forward-forward (FF) or tail-to-tail (RF) primers for the verifications of inversions and duplications.

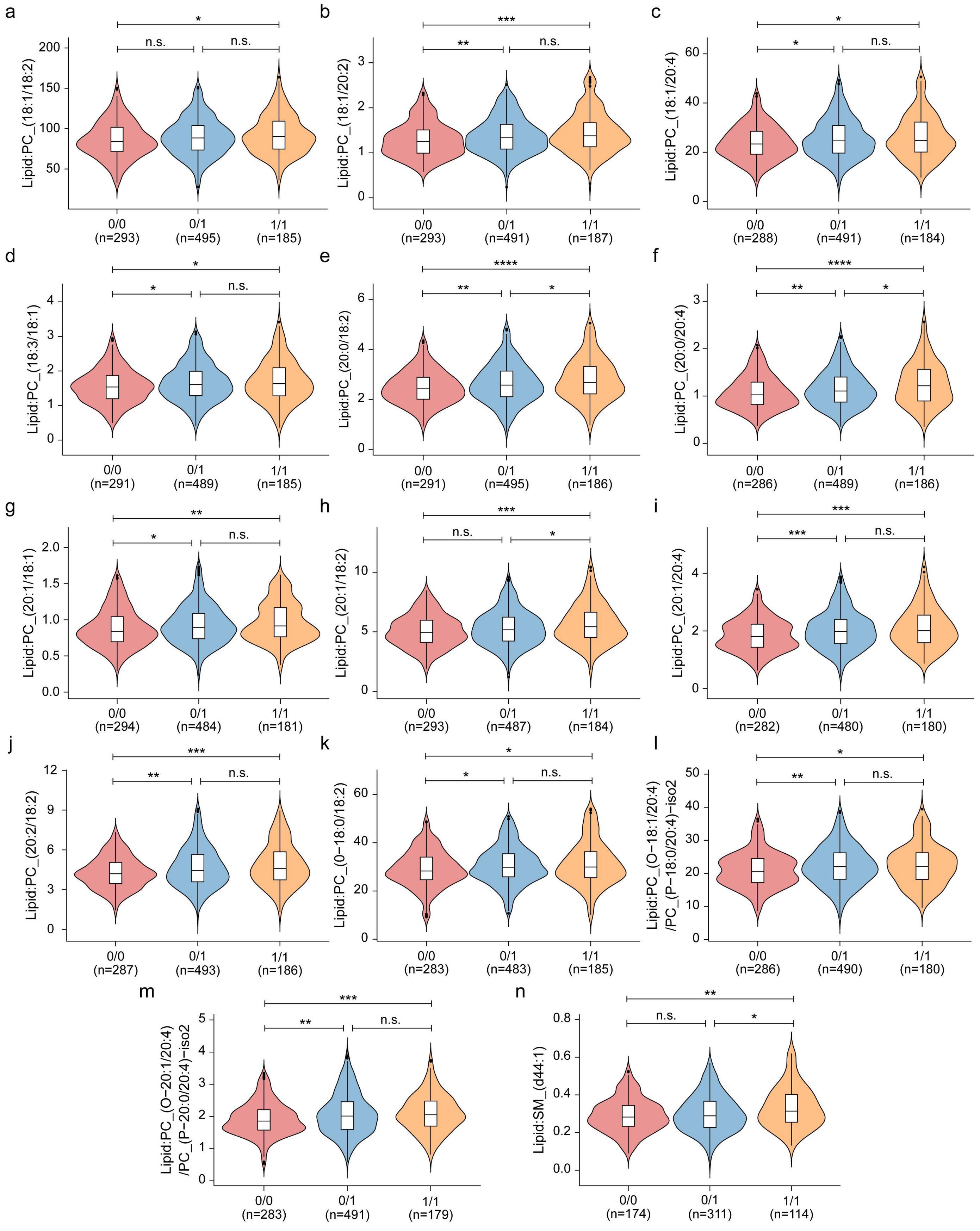


**Fig. S3:** Significant increases of 13 phosphatidylcholines (**a**, Lipid: PC_(18:1/18:2); **b**, Lipid: PC_(18:1/20:2). **c**, Lipid: PC_(18:1/20:4). **d**, Lipid: PC_(18:3/18:1). **e**, Lipid: PC_(20:0/18:2). **f**, Lipid: PC_(20:0/20:4). **g**, Lipid: PC_(20:1/18:1). **h**, Lipid: PC_(20:1/18:2). **i**, Lipid: PC_(20:1/20:4). **j**, Lipid: PC_(20:2/18:2). **k**, Lipid: PC_(0−18:0/18:2). **l**, Lipid: PC_(O−18:1/20:4)/PC_(P−18:0/20:4)−iso2 and **m**, Lipid: PC_(O−20:1/20:4)/PC_(P−20:0/20:4)−iso2) and sphingomyelin (**n**, Lipid: SM_(d44:1)) were observed in SV carriers than the in non-carriers. X-axes indicate the samples of different genotypes and Y-axes indicate the levels of 13 phosphatidylcholines and sphingomyelin. *P* values were calculated by Kruskal-Wallis test for a-n. *p <0.05, **p <0.01, ***p <0.001, ****p <0.0001, n.s. non-significant.

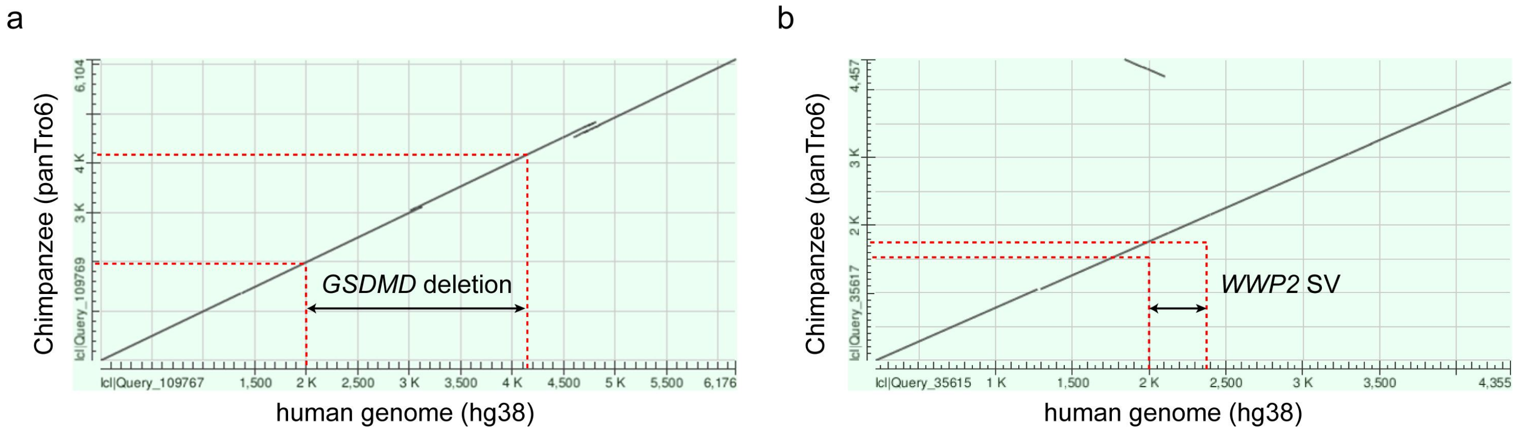


**Fig. S4:** Dot plots showing the alignment of the human and chimpanzee reference genomes at the *GSDMD* (**a**) and *WWP2* (**b**) loci using BLASTN. The marked regions are chr8:143,551,896–143,554,071 (GRCh38) at *GSDMD* locus and chr16:69,820,428–69,820,783 (GRCh38) at *WWP2* locus, respectively.

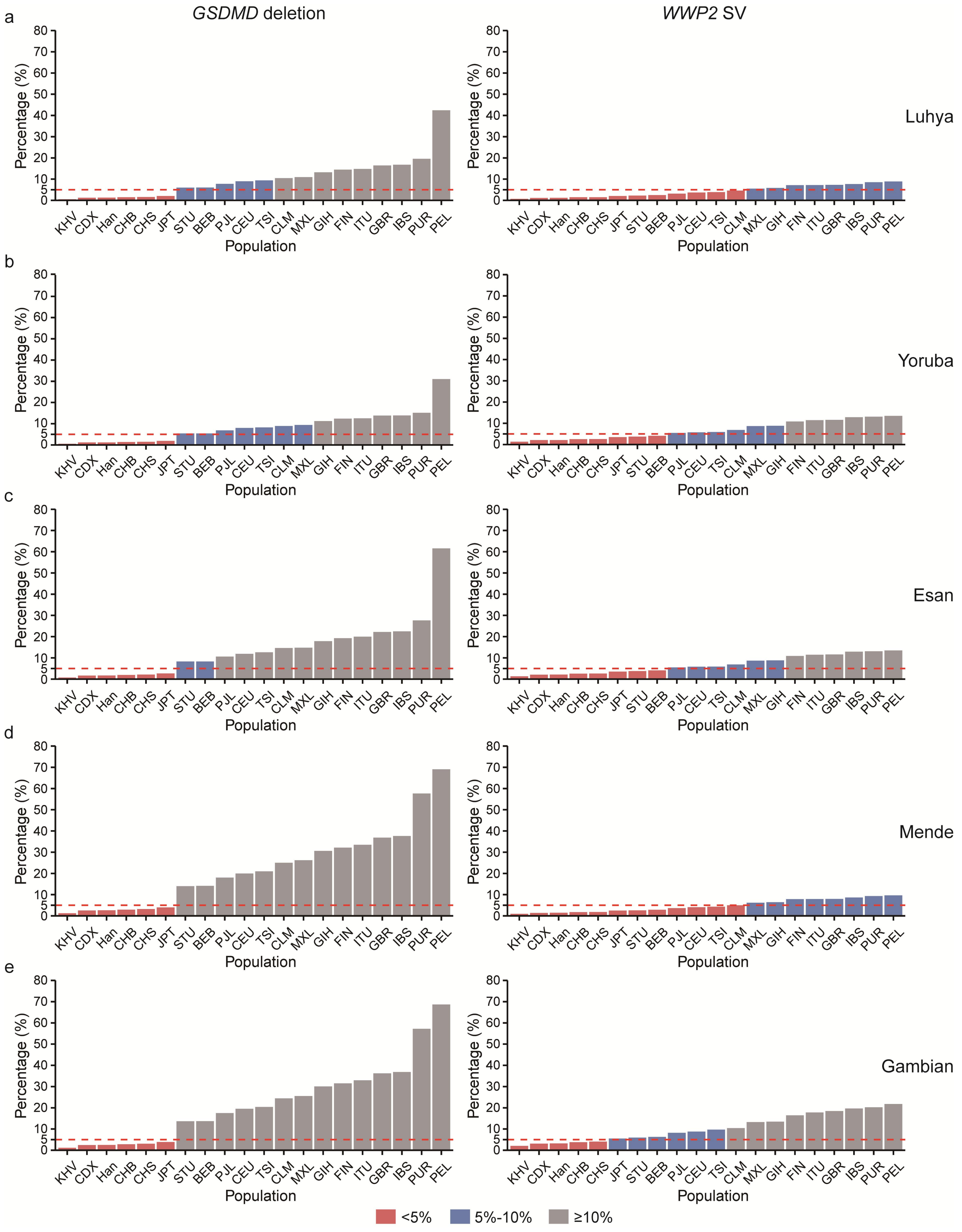


**Fig. S5:** Of the 102,119 structural variants (SVs) analyzed, the SVs at the *GSDMD* and *WWP2* loci exhibit among the highest levels of genetic differentiation, measured by $F_{ST}$ (the fixation index), between African and non-African populations. $F_{ST}$ was calculated between five African populations and 19 non-African populations from the 1000 Genomes Project, as well as an independent Han Chinese cohort (referred as "Han") from the present study. For the calculations, 50 samples were randomly selected to represent each population. The X-axis shows the names of the non-African populations analyzed, while the Y-axis shows the ranking percentile of the *GSDMD* and *WWP2* structural variants (SVs) based on genetic differentiation ($F_{ST}$) between African and non-African groups across 102,119 total SVs. Red, blue and gray bars denote percentile ranks below 5%, between 5-10%, and greater than or equal to 10%, respectively. The red dashed line indicates the $F_{ST}$ ranking cutoff at the 5th percentile.

**a**, $F_{ST}$ ranking of SVs at *GSDMD* and *WWP2* loci between Luhya and each of non-African populations. **b**, $F_{ST}$ ranking of SVs at *GSDMD* and *WWP2* loci between Yoruba and each of non-African populations. **c**, $F_{ST}$ ranking of SVs at *GSDMD* and *WWP2* loci between Esan and each of non-African populations. **d**, $F_{ST}$ ranking of SVs at *GSDMD* and *WWP2* loci between Mende and each of non-African populations. **e**, $F_{ST}$ ranking of SVs at *GSDMD* and *WWP2* loci between Gambian and each of non-African populations.

IBS: Iberian populations in Spain; GBR: British in England and Scotland; TSI: Toscani in Italy; FIN: Finnish in Finland; PJL: Punjabi in Lahore, Pakistan; GIH: Gujarati Indians in Houston, TX; ITU: Indian Telugu in the UK; STU: Sri Lankan Tamil in the UK; BEB: Bengali in Bangladesh; CDX: Chinese Dai in Xishuangbanna, China; CHS: Han Chinese South; CHB: Han Chinese in Beijing, China; JPT: Japanese in Tokyo, Japan; KHV: Kinh in Ho Chi Minh City, Vietnam; CEU: Utah residents (CEPH) with Northern and Western European; MXL: Mexican Ancestry in Los Angeles, California; PUR: Puerto Rican in Puerto Rico; CLM: Colombian in Medellin, Colombia; PEL: Peruvian in Lima, Peru.

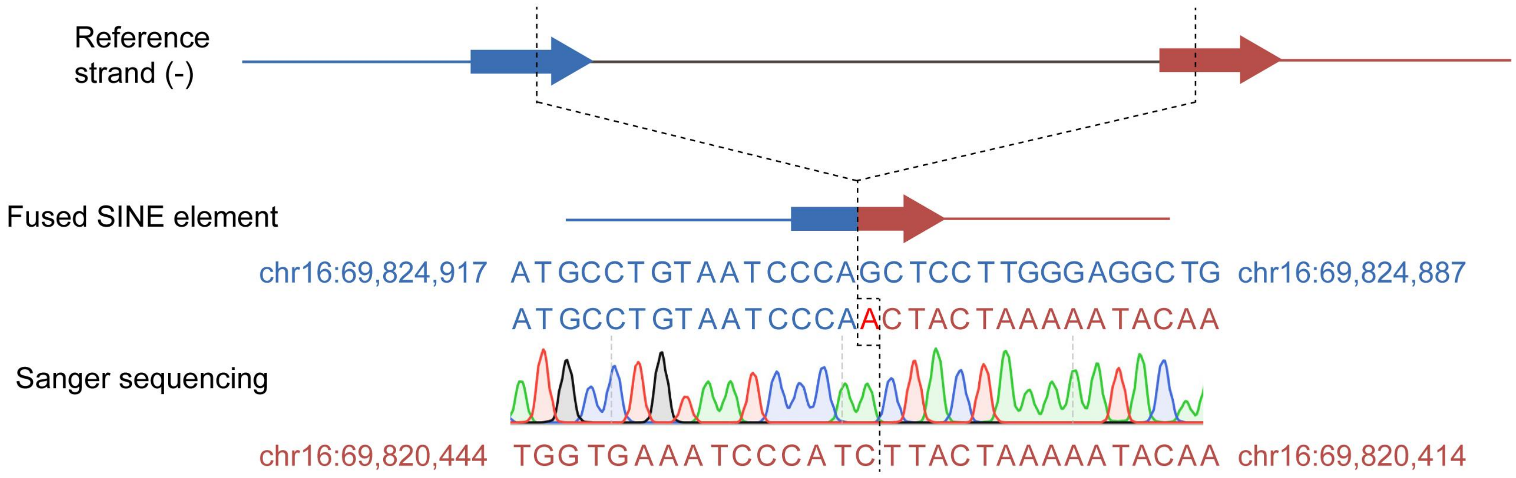


**Fig. S6:** Schematic of the SINE fusion event at the *WWP2* locus.

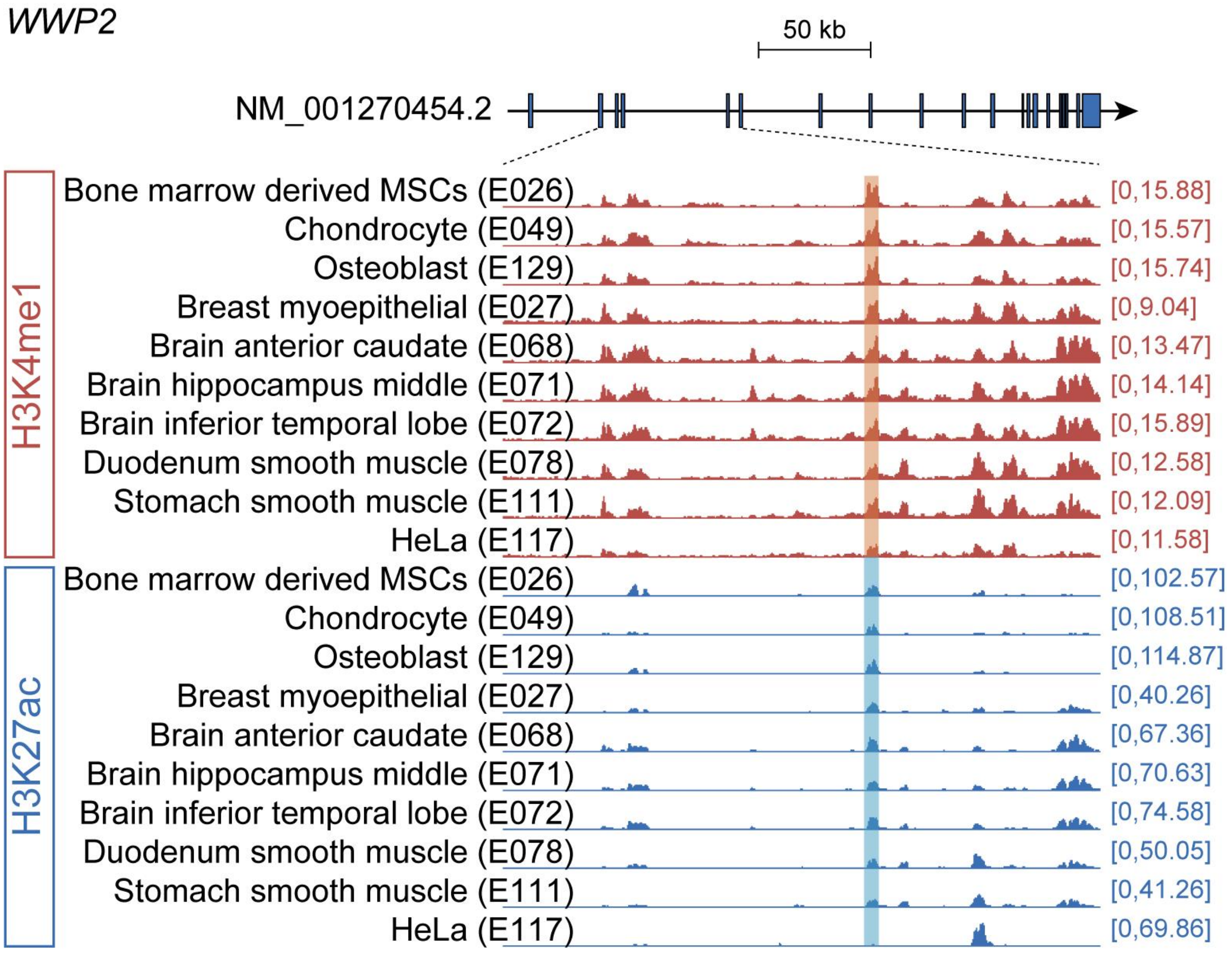


**Fig. S7:** The deletion in the fourth intron of *WWP2* (NM_001270454.2) eliminates a region that is enriched of histone modification signals. H3K4me1 and H3K27ac ChIP-seq tracks for Bone marrow derived MSCs (E026), Mesenchymal stem cell derived chondrocyte (E049), Osteoblast (E129), Breast myoepithelial (E027), Brain anterior caudate (E068), Brain hippocampus middle (E071), Brain inferior temporal lobe (E072), Duodenum smooth muscle (E078), Stomach smooth muscle (E111), Hela-S3 cervical carcinoma (E117) from the Roadmap Epigenomics dataset were visualized in the UCSC Genome Browser.

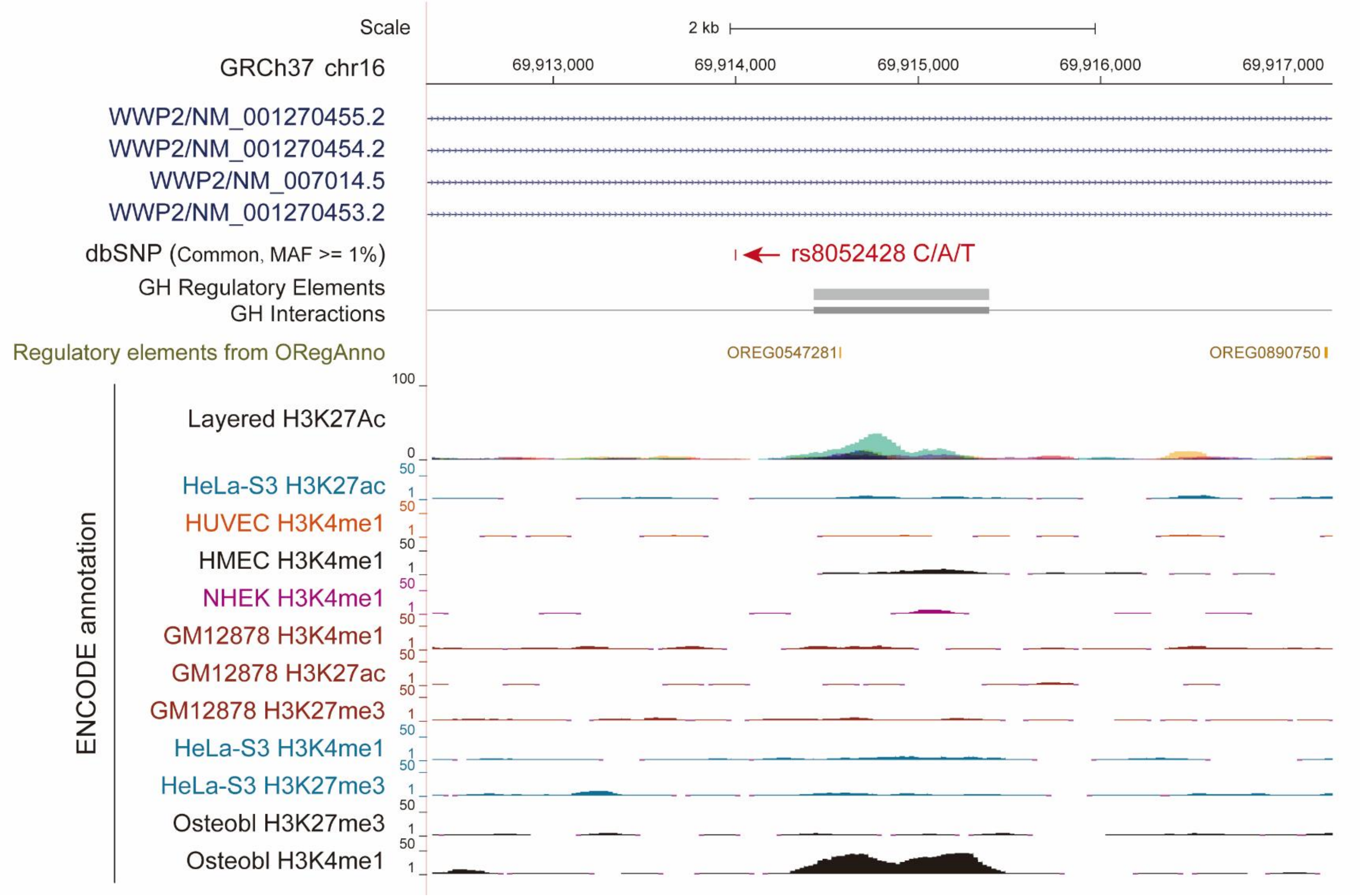


**Fig. S8:** There is no enhancer annotation surrounding rs8052428 (indicated by the red arrow), which is in linkage disequilibrium with the SV at the *WWP2* locus. The visualized annotation information includes regulatory elements from ORegAnno, enhancer annotations from the GeneHancer database, layered H3K27ac histone mark tracks across 7 cell lines (GM12878, H1-hESC, HSMM, HUVEC, K562, NHEK and NHLF), as well as H3K4me1, H3K27me3, and H3K27ac ChIP-seq tracks for HUVEC, Hela-S3, HMEC, NHEK, Osteoblasts, and GM12878 cell types from the ENCODE dataset. All annotations were visualized using the UCSC Genome Browser.

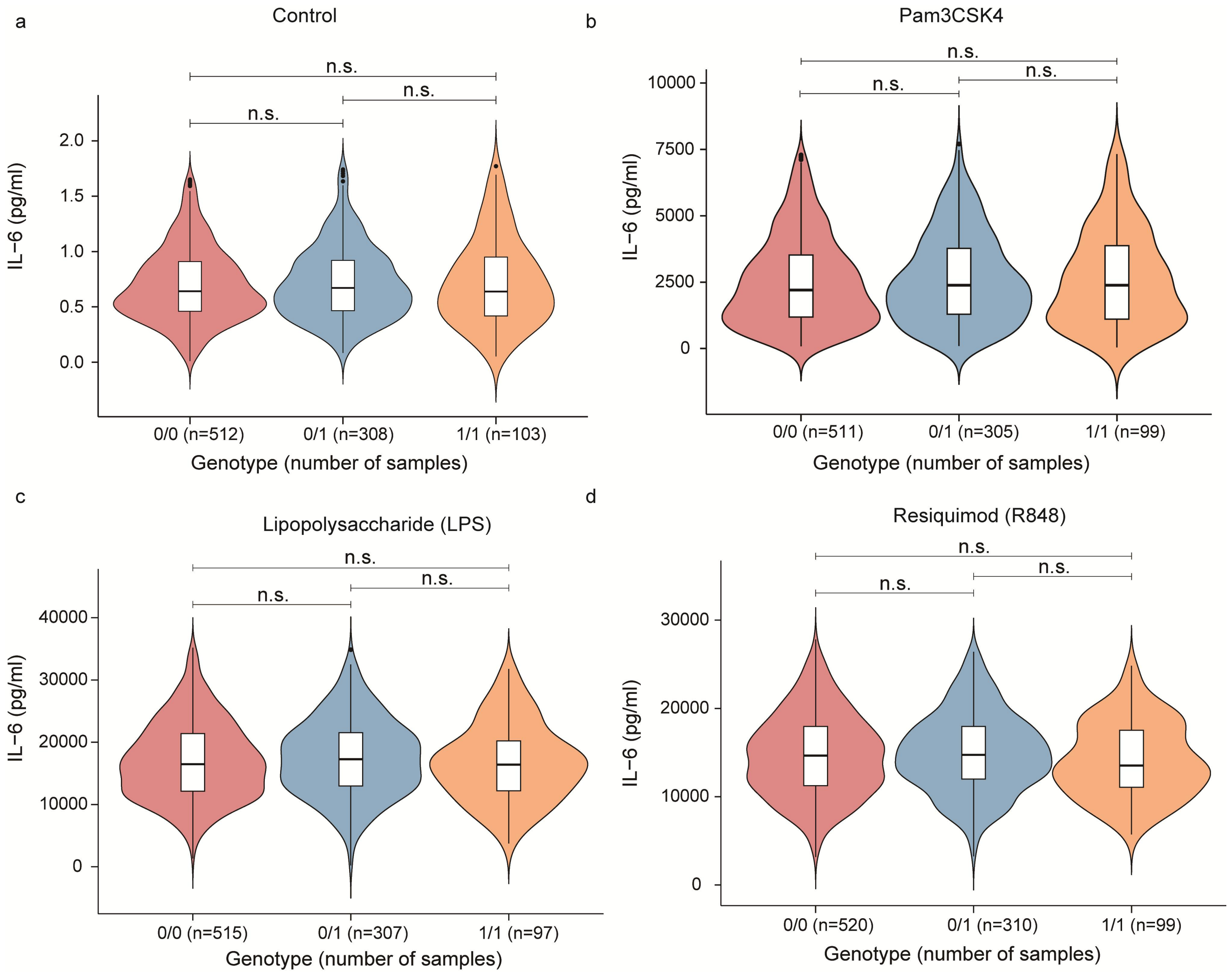


**Fig. S9:** No significant differences in IL-6 levels were observed between the carriers and non-carriers of the SV at the *WWP2* locus when no stimulus (negative control, **a**) or stimulated with Pam3CSK4 (**b**), lipopolysaccharide (LPS, **c**), and resiquimod (R848, **d**). X- and Y-axes indicate the samples of different genotypes and the level of IL-6, respectively. *P* values were calculated by Kruskal-Wallis test for **a-d**. n.s. non-significant.

**Separate Excel file**

Tables S1 to S9
